# BNT162b2 Vaccine Induces Divergent B cell responses to SARS-CoV-2 S1 and S2

**DOI:** 10.1101/2021.07.20.21260822

**Authors:** R. Camille Brewer, Nitya S. Ramadoss, Lauren J. Lahey, William H. Robinson, Tobias V. Lanz

**Author notes:** equal contributions.

## Abstract

The first ever messenger RNA (mRNA) vaccines received emergency approvals in December 2020 and are highly protective against SARS-CoV-2^1–3^. However, the contribution of each dose to the generation of antibodies against SARS-CoV-2 spike (S) protein and the degree of protection against novel variants, including delta, warrant further study. Here, we investigated the B cell response to the BNT162b2 vaccine by integrating repertoire analysis with single-cell transcriptomics of B cells from serial blood collections pre- and post-vaccination. The first vaccine dose elicits highly mutated IgA**^+^** plasmablasts against the S protein subunit S2 at day 7, suggestive of recall of a memory B cell response generated by prior infections with heterologous coronaviruses. On day 21, we observed minimally-mutated IgG**^+^** activated switched memory B cells targeting the receptor binding domain (RBD) of the S protein, likely representing a primary response derived from naïve B cells. The B cell response against RBD is specifically boosted by the second vaccine dose, and encodes antibodies that potently neutralize SARS-CoV-2 pseudovirus and partially neutralize novel variants, including delta. These results demonstrate that the first vaccine dose activates a non-neutralizing recall response predominantly targeting S2, while the second vaccine dose is vital to boosting neutralizing anti-S1 RBD B cell responses.

---

BNT162b2 is one of the two first widely available vaccines that are based on lipid nanoparticle delivery of modified mRNA and is dependent on the host cells for translation and cell surface expression of the SARS-CoV-2 spike (S) protein^4^. S consists of two subunits, S1 and S2. The S1 subunit contains the receptor binding domain (RBD) that binds human ACE2 and initiates viral cell entry, while S2 mediates viral cell membrane fusion^5, 6^. RBD is the target of most neutralizing antibodies found in COVID-19 patients^7^. Serological and repertoire studies from COVID-19 patients have characterized neutralizing antibodies to S protein^1, 2, 8, 9^. However, the cellular processes that lead to potent neutralizing antibodies in response to mRNA vaccines are not fully characterized and to what degree these antibodies protect against novel variants remains unclear.

## Single-cell sequencing of B cells

Nine healthy individuals were included in this study, all naive to prior SARS-CoV-2 infection (Extended Data Table 1). One individual contracted symptomatic and PCR-positive COVID-19 eight weeks after the second vaccine. Magnetically enriched peripheral blood B cells were investigated by droplet-based single-cell sequencing prior to the first vaccination (D0), as well as 7-9 days (D7), 21-23 days (D21), and 28 days (D28) after the first vaccination (Fig. 1a). All individuals received their second dose on D21. In addition, SARS-CoV-2 S-specific B cells were labeled with S1, S2, and RBD tetramers conjugated to fluorochromes and DNA-barcodes, and FACS-sorted before sequencing (Fig. 1a). A total of 131,138 B cells (∼3600 per sample) were included in the global B cell transcriptomic analysis. Dimensionality Reduction by Uniform Manifold Approximation and Projection (UMAP)^10^ and graph-based clustering distinguished nine B cell populations present in all individuals at all four timepoints, including the canonical naïve B cell, unswitched memory (UM) and switched memory (SM) B cell, and plasmablast (PB) clusters (Fig. 1b, Extended Data Fig. 1a-b). Naive B cells and SM were further categorized into resting and activated populations by *CD24* and *CD86* gene expression (Extended Data Fig. 1c-e)^11^. Cluster assignments are supported by isotype proportions and mutation frequencies (Extended Data Fig. 1f-g).

**Fig. 1:**
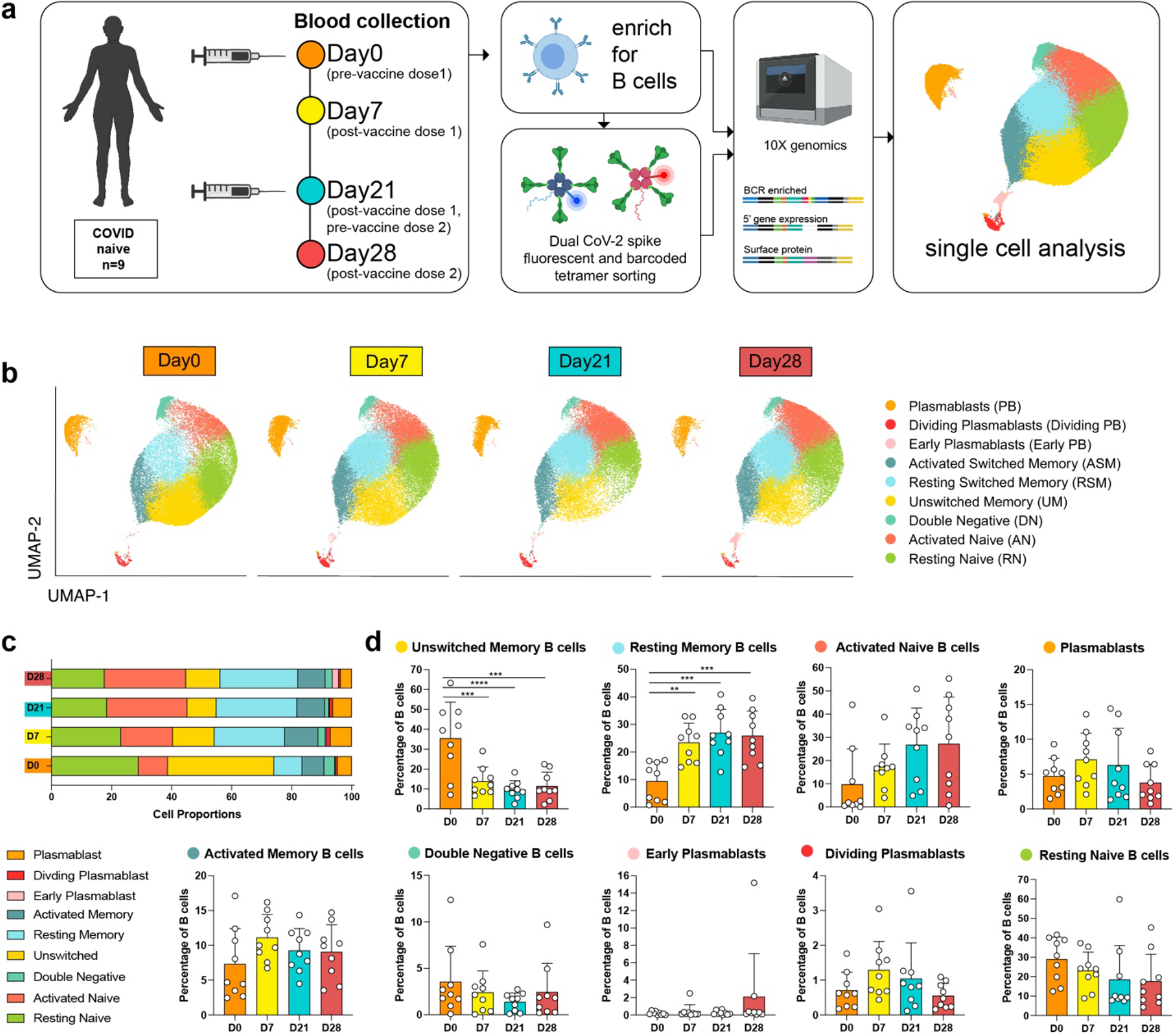
Single cell transcriptomic analysis of the B cell response following BNT162b2 vaccination. **a**, Outline of experimental approach. **b**, UMAP visualization of single-cell B cell transcriptome sequencing data of all individuals at four timepoints. B cell cluster assignments are based on gene expression and cell surface expression (CITE-seq) of canonical B cell markers. **c**, Mean percentages of B cell subtypes shown in (**b**). **d**, Individual and mean percentages of B cell types in (**b**) at four timepoints. *n*=9 individuals at all timepoints. Individual values, means and standard deviations are shown. \*\**P* <0.01, \*\*\**P* <0.001, \*\*\*\**P* <0.0001 according to two-tailed one-way ANOVA test in comparison to the first timepoint.

## Unswitched memory (UM) differentiation into resting memory B cells (RSM) in response to vaccination

To evaluate changes in the B cell transcriptome in response to the mRNA vaccine, B cell subtypes were followed over time (Fig. 1b-d). Strikingly, UM decreased after the initial vaccination, while RSM expanded (Fig. 1c,d). To track the diminishing UM, we followed clonally expanded UM from D0 to the post-vaccination timepoints by their clonal B cell receptor (BCR) sequences (Fig. 2a). Pre-vaccine UM develop into RSM in response to vaccination, with no significant change in mutation load (Fig. 2a, Extended Data Fig. 2a-b), suggesting differentiation without germinal center (GC) maturation. Clonal UM and RSM have little connectivity to PB or activated switched memory (ASM) B cell clusters, indicating activation and differentiation of UM is independent of the antibody response against SARS-CoV-2 (Fig. 2a, Extended Data Fig. 2c). Pathway enrichment analysis of UM and RSM determined significant changes in leukocyte activation, cytokine signaling, and B cell signaling pathways post vaccination (Extended Data Fig. 3a,b). Accordingly, we observed increased expression of B cell signaling and activation genes in UM (Fig. 2b). In contrast, we observed downregulation of the same genes in RSM after vaccination (Fig. 2c). However, upregulation of CD83 and CD69 in RSM indicates recent activation (Extended Data Fig. 3d,e)^12, 13^. Together, this suggests that vaccination activates UM and induces class-switching and differentiation into RSM, independent of the GC. RSM, once differentiated, downregulate their activation levels.

**Fig. 2:**
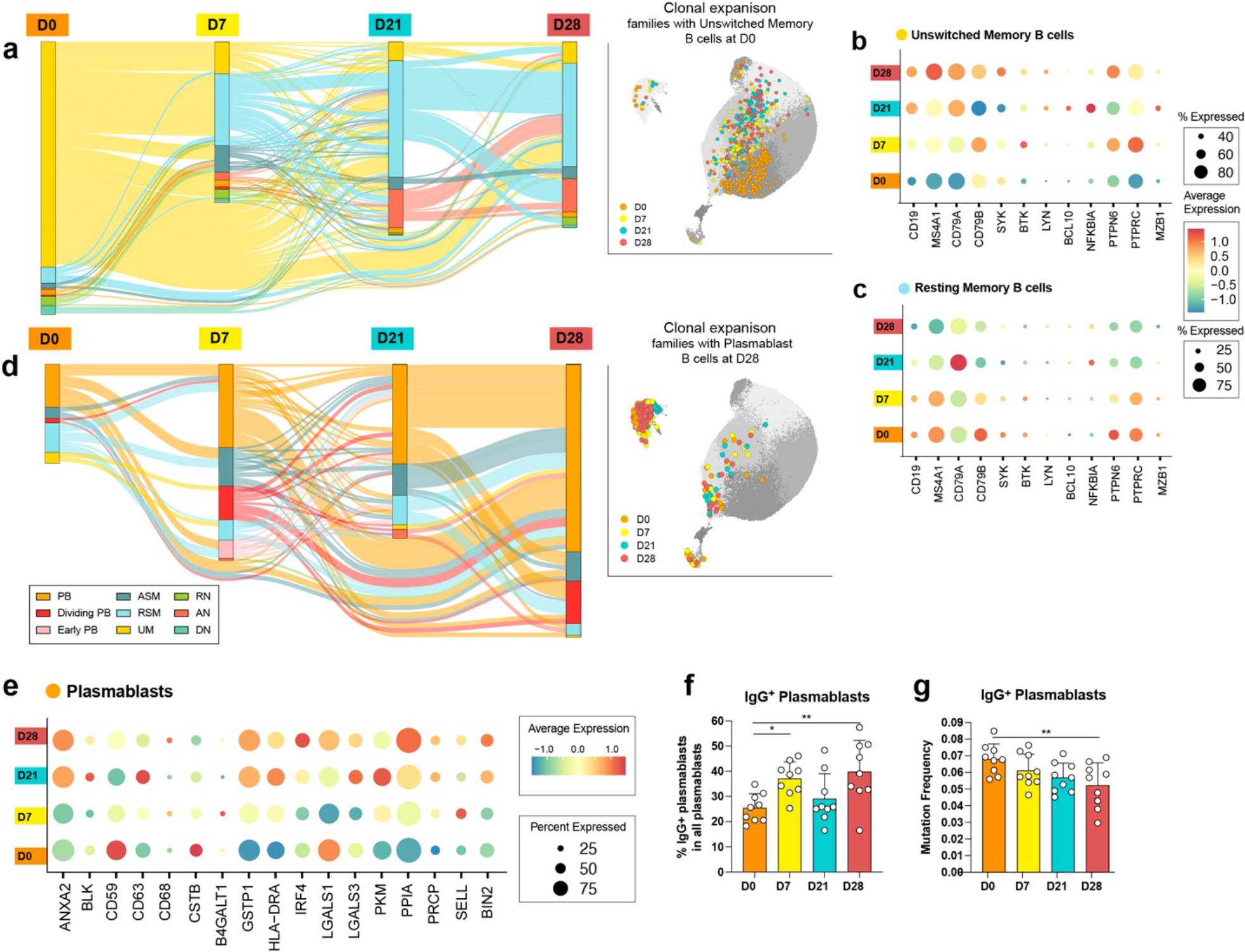
Unswitched memory B cell differentiation and expansion of IgG^+^ plasmablasts in response to vaccination. **a**, Alluvial plot of B cell trajectories (left) across timepoints and UMAP projection (right) of clonally expanded B cells that are related to at least one UM at D0. Clonal expansions included have ≥3 members at ≥2 timepoints. **b-c**, Gene expression in UM (**b**) and RSM (**c**) of genes involved in B cell signaling and activation. **d**, Alluvial plot of B cell trajectories (left) and UMAP projection (right) of clonally expanded B cells that are related to at least one PB at D28. Clonal expansions included have ≥3 members at ≥2 timepoints. **e**, Gene expressions of activation markers in PB. **f**, Percentage of IgG^+^ PB per individual (n=9), at four timepoints. **g**, Average mutation frequency of IgG^+^ PB per individual (*n*=9) at four timepoints. Individual values, means and standard deviations are shown. \**P* < 0.05; or \*\**P* <0.01 according to two-tailed one-way ANOVA test in comparison to the first timepoint.

**Fig. 3:**
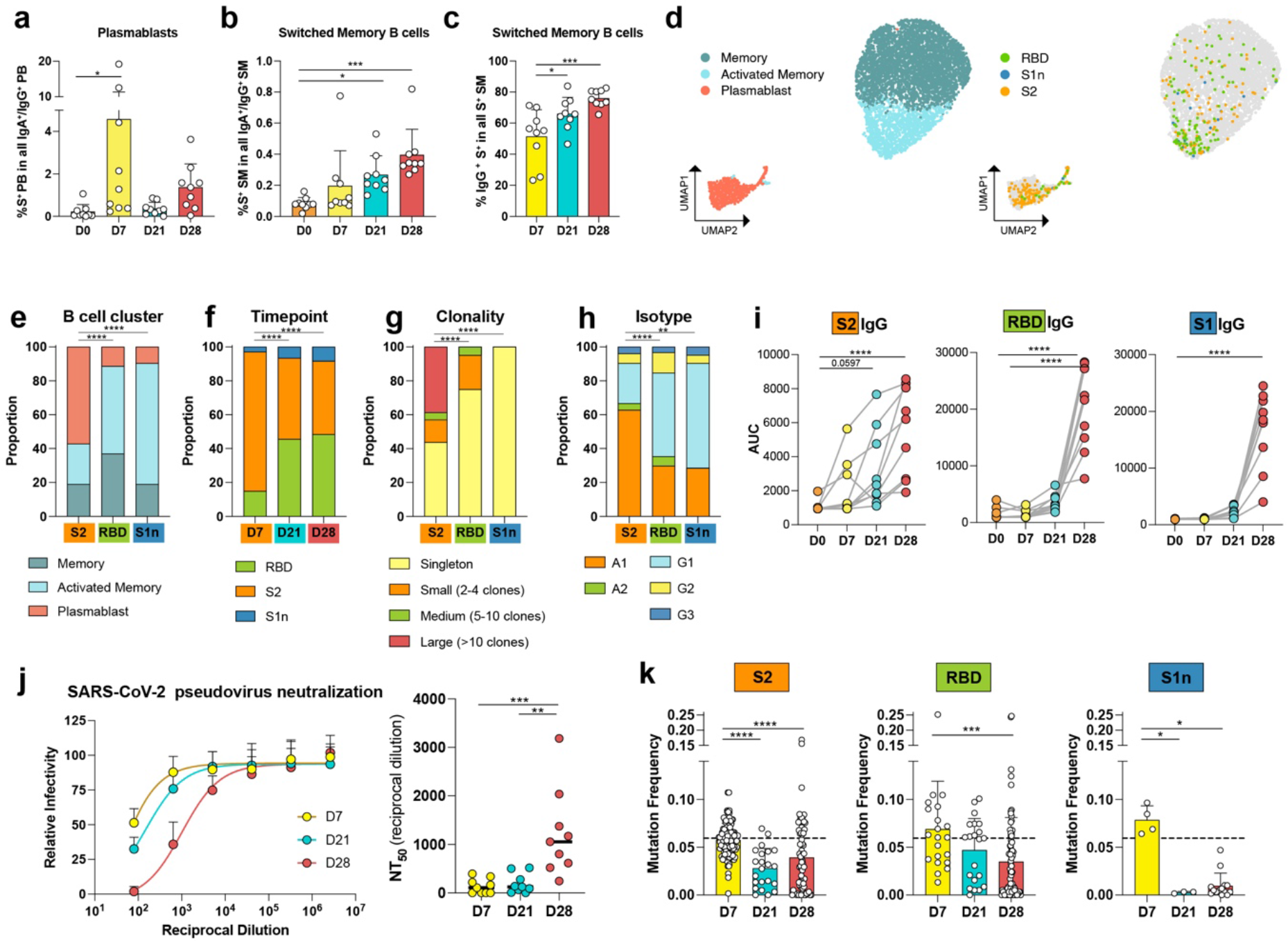
Vaccination induces a IgA^+^ anti-S2 response on day 7 followed by IgG^+^ RBD response on days 21 and 28. **a-c**, Quantification of flow cytometry data, showing percentages of **a**, anti-S PB, and **b**, anti-S SM in all B cells at four timepoints. **c**, Percentage of IgG^+^ anti-S SM in all anti-S SM at four timepoints. *n*=9 individuals per timepoint. Individual data points are averages of two independent experiments. **d**, UMAP visualization of demultiplexed transcriptomic data of anti-S^+^ and anti-S^-^ sorted B cells, showing cluster assignments of memory (blue), activated memory (turquoise), and PB (red) (left) and antigen-specificity to S2 (orange), RBD (green), and S1n (blue) (right). **e-h**, Proportions of sorted cells shown in (**d**), separated by antigen and by **e**, cluster distribution, **f**, timepoint, **g**, clonality, and **h**, isotype. **i**, Plasma IgG levels against S2 (left), RBD (center), and S1 (right) for n=9 individuals at four timepoints. AUC for plasma dilutions are shown. **j**, Plasma neutralization curves (left) and quantification (NT50) of neutralization titers (right) of n=9 individuals at timepoints post vaccination. **k**, V-gene mutation frequencies (number of mutations per length of V-gene) of B cells specific for S2 (left), RBD (center), and S1n (right) at timepoints post vaccination. Dashed line indicates the average mutation frequency of sorted antigen negative B cells. Individual data points represent single B cells from n=9 individuals. Means and standard deviations are shown. *P < 0.05, \*\**P* <0.01, \*\*\**P* <0.001, \*\*\*\**P* <0.0001 according to two-tailed one-way ANOVA test in comparison to the first timepoint displayed and **g-j**, chi-square test.

## PB and ASM recruitment in response to vaccination

PB are short-lived, activated B cells that expand in response to antigen stimulation and secrete large amounts of antibodies^14^. In contrast to strong PB expansions in other vaccination settings^15^, overall PB numbers in our study only slightly increase post SARS-CoV-2 vaccination (Fig. 1d, Extended Data Fig. 4a). To identify B cell populations directly involved in the antigen-specific response, we focused on clonal B cell expansions that include PB at D28 (Fig. 2d). Based on shared BCR sequences, we found strong connectivity between the PB, dividing PB, and ASM subsets (Fig. 2d, Extended Data Fig. 2c). Genes associated with leukocyte activation and protein processing are enriched in PB, consistent with their function as antibody producers (Extended Data Fig. 3c). Enrichment of genes associated with SARS-CoV-2 infection at D21 suggests that recognition of viral proteins trigger similar pathways in immunization and infection (Extended Data Fig. 3c). Overall, the majority of genes associated with B cell activation increase over time in response to vaccination (Fig. 2e).

**Fig. 4:**
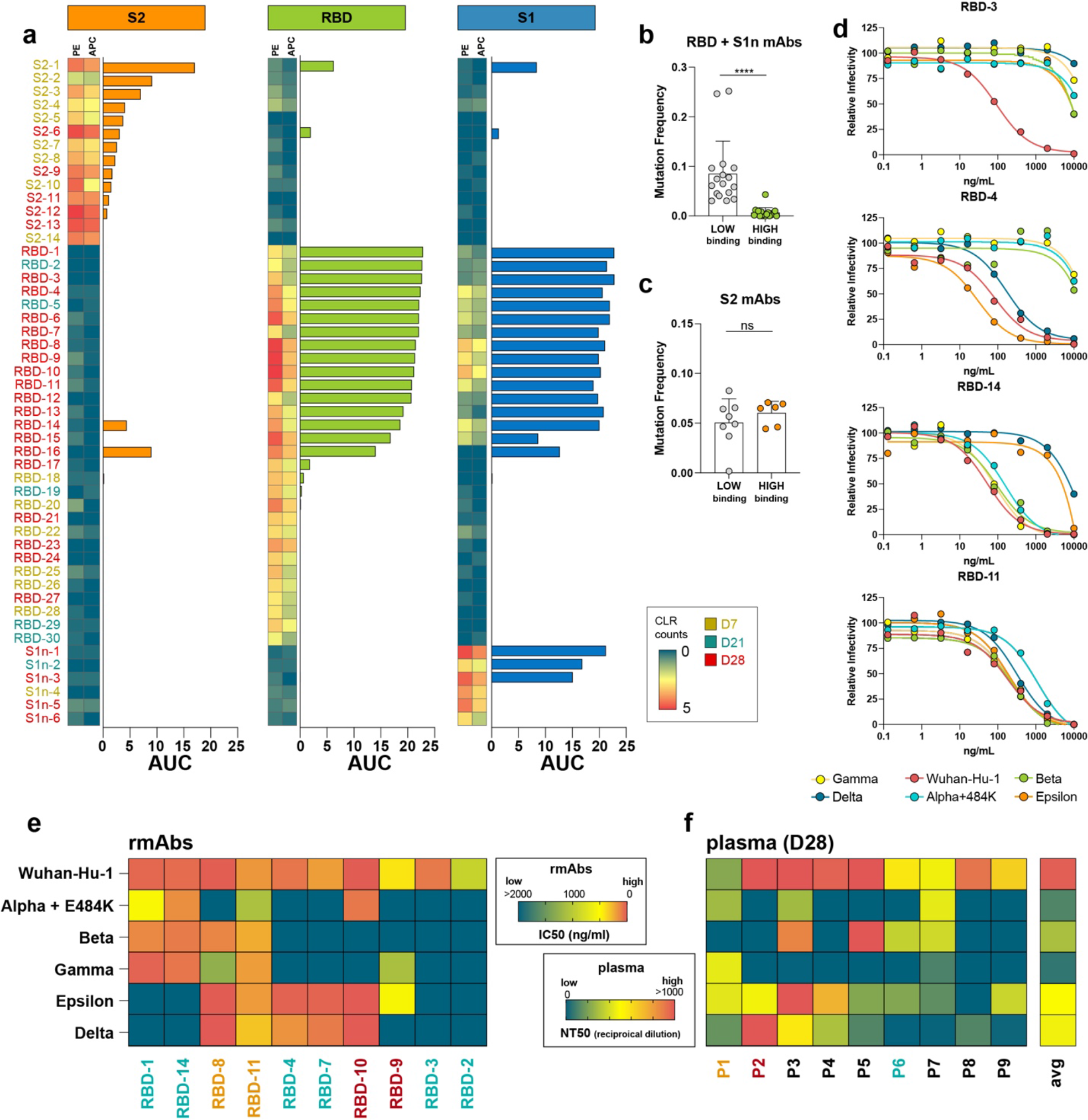
Neutralization of SARS-CoV-2 pseudovirus and variants by BNT162b2-induced antibodies. **a**, recombinant mAbs derived from BNT162b2-induced B cells bind S2, S1 RBD, and S1.Two-column heatmaps show barcoded tetramer sequencing data (PE and APC labeled) represented as centered log ratio transformed (CLR) counts. Bar graphs show binding of rmAbs generated from the corresponding B cells, represented as area under the curve (AUC) of serial dilutions, measured by ELISA. Threshold, represented as 0, was set to the average binding to BSA plus three times the standard deviation of background binding to bovine serum albumin (BSA). **b-c**, Mutation frequencies (number of mutations per length of V-gene) of **b**, anti-RBD and S1, and **c**, anti-S2 binding rmAbs from (**a**). High binding rmAbs are defined as positive for double-barcoded tetramer binding and AUC >3. **d**, Neutralization curves for rmAbs RBD-3, RBD-4, RBD-14, and RBD11, blocking Wuhan-Hu-1 pseudovirus and variants Alpha+E484K, beta, gamma, epsilon, and delta. **e-f**, Heatmaps indicating **e**, IC50 (ng/ml) for each rmAb (**e**) and **f**, NT50 (reciprocal dilution) of D28 plasma of n=9 individuals to the indicated pseudovirus variants. \*\*\*\**P* <0.0001 according to unpaired two-tailed t-test.

In health, a high proportion of peripheral blood PB are IgA^+^ due to ongoing immune responses at the mucosal barriers. IgG^+^ PB of non-mucosal origin increase during systemic infection and vaccination^16, 17^. Congruently, we observed an increase in IgG^+^ PB one week after the first and second vaccination, respectively (Fig. 2f, Extended Data Fig. 4b-e), with strongest expansions in the inflammatory subtypes IgG1 and IgG3 (Extended Data Fig. 4c). The average mutation frequencies of IgG^+^ PB decrease over time, indicative of an influx of cells into PB from the naïve B cell pool rather than from memory B cells (Fig. 2g). Together, our results show that clonal PB and ASM are closely connected, and new, low mutation IgG^+^ PB are recruited in response to vaccination.

## S-specific B cells expand in response to vaccination

To further characterize the S-specific B cell response to vaccination, we labeled B cells with fluorescently labeled S1, S2, and RBD antigen tetramers, each with unique barcodes, and FACS-sorted tetramer-specific and non-specific IgA^+^/G^+^ PB and IgA^+^/G^+^ SM (Extended Data Fig. 5a). Antigen-specificity of each single B cell could be determined by their barcode after demultiplexing (Extended Data Fig. 6a-d). In contrast to the global PB population (Fig. 1d), and in line with our observation on the IgG^+^ PB population (Fig. 2f), S-specific IgA^+^/G^+^ PB strongly expand one week after the initial vaccination and to a lesser degree after the second vaccination (Fig. 3a, Extended Data Fig. 5b). The S-specific IgA^+^/G^+^ SM response increases over time from D7 to D28 (Fig. 3b, Extended Data Fig. 5d). Both S-specific populations, SM and PB, shift from IgA^+^ to IgG^+^ over time (Fig. 3c, Extended Data Fig. 5c,e).

## Differential B cell response to S1 and S2

Vaccine studies have shown that antibody responses against distinct epitopes contribute differentially to immune protection^18^. Here, we investigated how the antibody response to each subunit of S varies within B cell subsets and over time. UMAP-analysis revealed that S2-specific B cells are predominantly PB and appear in large numbers as early as D7 (Fig. 3d-f). In contrast, the majority of RBD-specific B cells and B cells that bind S1 but not RBD (S1n) are ASM (Fig. 3d,e, Extended Data Fig. 5f). Additionally, the RBD and S1n-specific B cells need longer to develop, accounting for less than 20% of S-specific B cells on D7 and over 50% by D28 (Fig. 3d-f,). The S2-specific PB response is highly clonal and IgA1^+^ dominated, while RBD and S1n-specific ASM use predominantly IgG1, and are less clonally expanded than S2 PB (Fig. 3g,h). The rapid recall of S2 and delayed S1 response indicates that the S2 B cell response is a secondary response, while the B cell response to RBD is a primary response.

The early S2 PB response is echoed by the early development of anti-S2 plasma antibody titers, which start leveling off at D21. Strikingly, RBD and S1 IgG and IgA titers remain low until day 28, one week after the second vaccination (Fig. 3i, Extended Data Fig. 7a-d). Accordingly, plasma neutralization of SARS-CoV-2 Wuhan-Hu-1 pseudovirus is significantly boosted after the second vaccination (Fig. 3j). As primarily anti-RBD antibodies have the potential to block viral entry into the cell, this differentiation is critical and highlights the importance of the second injection for a protective anti-RBD response. Notably, it is overlooked in studies that focus solely on the anti-S antibody response.

Interestingly, one study participant contracted COVID-19 eight weeks after the second vaccination. The individual exhibited the lowest IgG and IgA titers for RBD and the lowest plasma neutralization potency following both vaccine doses in the tested cohort (Extended Data Fig. 8a-c). While this individual did produce B cells with high-affinity neutralizing BCR sequences (P1 in Fig. 4e,f), antibody secretion seemingly did not occur on sufficient levels. Antibody titers did not increase significantly even two weeks after the infection (Extended Data Fig. 8d), suggesting that low antibody titers in this individual are not confined to the mRNA vaccine.

## Influx of minimally-mutated anti-RBD B cells

During antigen-specific B cell responses, BCR sequences tend to accumulate mutations during affinity maturation^19^. In contrast, neutralizing antibodies from COVID-19 patients show characteristically low mutation rates, indicating recruitment of naïve B cells to the GC in response to a novel antigen that has little structural overlap with previously encountered pathogens^20^. In our study, D7 B cells specific for any of the three antigens showed mutation frequencies similar to the antigen-negative sorted B cells, suggesting that this initial response stems directly from the memory B cell pool (Fig. 3k). At this early timepoint, S2-specific B cells predominate over RBD and S1n-specific B cells, providing further evidence that the rapid memory recall response is more effective for S2 than S1 (Fig. 3f, k). Consistent with this observation, S2 is more conserved among human pathogenic coronaviruses than S1^21, 22^. Strikingly, at timepoints D21 and D28, with the influx of S1-specific B cells, mutation frequencies decrease (Fig. 3k). Together, these results indicate new recruitment from the naïve B cell pool and short GC maturation in response to the vaccine.

## Vaccine-induced neutralization of variants

To test if these newly-recruited, minimally mutated anti-S antibodies are better at binding the S subunits than the early, highly mutated anti-S antibodies recruited from the memory B cell pool, we expressed 50 representative BCR sequences as recombinant monoclonal antibodies (rmAbs) — 14 from S2-specific, 30 from S1/RBD-specific, and 6 from S1n-specific B cells (Extended Data Table 2). Of those selected antibodies, 8 rmAbs bound S2, 15 rmAbs bound RBD, and 3 rmAbs bound S1 but not RBD, as measured by ELISA and bio-layer interferometry (Fig. 4a and Extended Data Fig. 9). Interestingly, when we compared low and high binding anti-S1 rmAbs (encompassing RBD and S1n) we found that high binding rmAbs show significantly lower mutation frequencies than low binding anti-S1 rmABs (Fig. 4b). In contrast, mutation frequencies of rmAbs against S2 did not differ significantly between low and high binders (Fig. 4c). Additionally, the S2 binders with the highest affinities were derived from D7 PB, while highest affinity antibodies against S1 and RBD stemmed from D21 and D28 (Fig. 4a). S2 is the more highly conserved subunit of S^22, 23^ To determine if the rmAbs were cross-reactive to other beta-coronaviruses, we tested the highest binders to RBD and S2 against the spike protein of four pathogenic beta-coronaviruses — HCoV-229E, HCoV-HKU1, HCoV-NL63, HCoV-OC43. Of note, nine of the 12 rmAbs against S2 cross-react with HCoV-OC43 spike protein. Conversely, none of the tested anti-RBD rmAbs are cross-reactive to non-SARS-CoV-2 coronaviruses (Extended Data Figure 10). These results corroborate that anti-S2 B cells originate from a recall memory response to heterologous coronaviruses, while in order to generate protective high-affinity anti-RBD antibodies, maturation of naïve cells to PB and ASM is required. While increased affinity correlating with low mutation frequency is counter-intuitive, it is characteristic for the B cell repertoire in COVID-19 patients^24, 25^, and likely caused by the novelty of S protein with little structural overlap to other pathogens, requiring strong recruitment from the naïve B cell pool.

The emergence of novel SARS-CoV-2 variants poses additional challenges to the management of the pandemic, and could jeopardize vaccine efficacy and the prospects of an expeditious return to normalcy. RBD mutations contribute significantly to immune escape. To evaluate the effects of variants on antibody neutralization, we selected ten high-affinity Wuhan-Hu-1-neutralizing anti-RBD rmAbs from three individuals (P1, P2, P6) with different levels of plasma neutralization against delta (Fig. 4d-f). Five different SARS-CoV-2 variants of concern were included (Extended Data Table 3) –– variants alpha+E484K (B.1.1.7+E484K; N501Y, E484K), beta (B.1.351; N501Y, K417N), and gamma (P.1; N501Y, K417T) share a similar set of RBD mutations, of which N501Y contributes most to immune escape^26^. A different mutation, L452R, is shared by delta (B.1.617.2; L452R, T478K) and epsilon (B.1.429; L452R), and also promotes escape from antibody neutralization^27^. Each variant can be neutralized by at least four of the ten rmAbs, highlighting the vaccine’s potency to generate at least partial protection to all variants (Fig. 4d,e, Extended Data Fig. 11). Antibody neutralization follows RBD mutations, as antibodies tend to block either alpha, beta, and gamma (N501Y), or delta and epsilon (L452R). Two antibodies (RBD-8 and RBD-11) were broadly neutralizing against all variants. Plasma neutralizations showed significantly lower potency against variants than against Wuhan-Hu-1 in all patients, while trending to a better neutralization of delta and epsilon than of alpha, beta, and gamma (Fig. 4f). Interestingly, while the plasma of individual P6 showed very limited neutralization efficacy to alpha, gamma, and delta, we show that P6 had potent neutralizing B cell clones against all variants. P6, therefore, will likely have an extended level of protection from a recall memory B cell response post vaccine, which is inaccessible to assessment by measuring plasma neutralization.

## Discussion

Our study provides a detailed characterization of the B cell response to the BNT162b2 mRNA vaccine on a single-cell level. Parsing the anti-S1 and S2 responses provides important insights into why the second vaccine dose is vital for protection. Our results demonstrate that the first vaccine dose activates a non-neutralizing recall response predominantly targeting epitopes in the S2 protein subunit, which is conserved across human-pathogenic coronaviruses^22, 23^, while the second vaccine dose is vital to boosting neutralizing B cell responses to S1 and RBD.

The vaccination induces two major shifts in distinct B cell subtypes: (i) activation of UM and their differentiation into RSM, and (ii) activation and expansion of S antigen-specific PB and ASM. UM are thought to develop independently of the GC reaction, and possess a polyreactive repertoire for rapid B cell responses^28^. UM are broadly activated early after vaccination, consistent with a natural or ‘innate-like’ B cell response. The unchanged mutation frequency post differentiation indicates a GC-independent response. Once switched, activation levels in RSM decrease, indicating that they are not further promoting an activated B cell response.

In contrast, the antigen-specific PB and ASM response is likely derived from GC-dependent processes. The first vaccination induces an IgA-dominant PB response against S2 with high mutation frequencies, which is cross-reactive to the human-pathogenic beta-coronaviruses OC43 and HKU1. Our data are consistent with a recall response of mucosal memory B cells that matured during prior pulmonary coronavirus infections.

After this initial response, we observed an influx of minimally mutated ASM on D21 and D28, which target S1 and RBD. We show that low mutation frequency corresponds to high affinity against RBD. mRNA vaccines have been shown to induce robust and prolonged GC reactions, with PB and SM persisting in GC for over three months^29, 30^. Despite their low mutation frequency, the delayed development and the switched phenotype of S1/RBD-specific PB and ASM indicate that they underwent GC maturation. High BCR affinity and a naïve phenotype foster preferential recruitment into GC^31–33^ and high affinity also promotes release from the GC as PB, plasma cells, or memory B cells^34^. High affinity of minimally mutated BCRs could therefore limit GC maturation to a relatively short time frame.

We utilized plasma samples and our low mutation RBD-binding rmAbs to characterize neutralization against several SARS-CoV-2 variants of concern. While we found a significant degree of immune escape, we also identified antibodies with neutralization potency against each variant. Importantly, we observed potent neutralization activity against the highly infectious delta variant, which has quickly become the dominant strain world-wide. Delta-neutralizing B cells were even found in an individual with very limited plasma neutralization potency, fueling hope that even individuals with low neutralizing titers can raise a recall memory response upon infection with delta.

Together, our study provides a detailed characterization of the blood B cell response to the BNT162b2 mRNA vaccine. Our data emphasize the importance of the second vaccine dose in inducing generation of anti-S1 RBD antibodies that contribute to neutralization of SARS-CoV-2 variants, including delta.

## Materials and methods

### Study design, sample collection, and storage

All studies were approved by the Institutional Review Board of Stanford University (IRB-3780), and the studies complied with the relevant ethical regulations. All participants provided written informed consent before participating in the study. Nine healthy individuals were enrolled in the study (Extended Data Table 1). All individuals had undergone routine RT-PCR testing prior to study. None of the participants had been previously diagnosed with SARS-CoV-2 infection. Blood samples were collected in heparin tubes (BD) at four different timepoints including pre-vaccination (D0), 7- to 9 days post initial vaccination (D7), on the day of and prior to the second dose (21- to 23 days post initial vaccination, D21), and 28 to 30 days after initial and 7-9 days after second vaccination (D28). Plasma samples were obtained after centrifugation, and stored at -80°C. Peripheral Blood Mononuclear Cells (PBMCs) were obtained by density gradient centrifugation over Ficoll PLUS media (Cytiva) and stored in cell freezing media (Thermo Fisher Scientific). Plasma samples were aliquoted and stored until use at -80°C.

### Generation of barcoded fluorescent antigen tetramers

Recombinant Avi-tag biotinylated SARS-CoV-2 S2 protein (Acro Biosystems, S2N-C52E8-25ug), SARS-CoV-2 RBD (Acro, SPD-C82E9-25ug), and SARS-CoV-2 S1 (Acro, S1N-C82E8-25ug) were mixed with barcoded, fluorescently labeled streptavidin (Biolegend) at a 4 to 1 molar ratios for 45 minutes while rotating. Excess biotin was added to saturate all streptavidin binding sites.

### Flow cytometry, cell sorting, and 10X sample preparation

PBMCs were thawed at 37C°, treated for 15min with DNAse and washed in complete RPMI. PBMCs were enriched for B cells using the EasySep Human Pan-B Cell Enrichment Kit (Stem Cell Technologies) according to the manufacturer instructions. B cell samples without antigen enrichment were stained with CD19, IgD, CD27, CD38 TotalSeq-C antibodies (all Biolegend). For antigen-sorted B cell samples, cells were stained with the following fluorescently labeled antibodies according to standard protocols: CD19, CD20, CD38 (all BD Biosciences), CD3, CD27, IgM, IgD, (all BioLegend), IgA (Miltenyi Biotec), Sytox blue (Thermo Fisher Scientific), and S-antigen tetramers (Extended Data Fig. 3a,b,d). Additionally, samples were labeled with TotalSeq-C hashtag 1-9 antibodies (Biolegend) for demultiplexing individual samples in downstream analysis. Single cells were sorted with a FACSAria II cell sorter (BD Biosciences) into cooled 1.5 ml tubes (BioRad). FlowJo Version 10.7.1 (BD Biosciences) and R version 3.6.1 was used for flow cytometry data analysis.

### Droplet-based single-cell sequencing

Using a Single Cell 5′ Library and Gel Bead Kit v1.1(10X Genomics, 1000165) and Next GEM Chip G Single Cell Kit (10X Genomics, 1000120), the cell suspension was loaded onto a Chromium single cell controller (10X Genomics) to generate single-cell gel beads in the emulsion (GEMs) according to the manufacturer’s protocol. Briefly, approximately 8,000 cells were added to each channel and approximately 4,000 target cells were recovered. Captured cells were lysed and the released RNA was barcoded through reverse transcription in individual GEMs. 5’ Gene expression (GEX) libraries, Single Cell V(D)J libraries (1000016), and Cell surface protein libraries were constructed according to manufacturer protocols. Library quality was assessed using a 2200 TapeStation (Agilent). The libraries were sequenced using an Illumina Novaseq6000 sequencer with a paired-end 150-bp (PE150) reading strategy (Novogene).

### Single cell RNA-seq data processing

Raw gene expression and cell surface matrices were generated for each sample by the Cell Ranger Pipeline (v.6.0.1) coupled with human reference version GRCh38. Briefly, gene expression analyses of single cells were conducted using the R package Seurat (v4.0.2) to perform data scaling, transformation, clustering, dimensionality reduction, differential expression analyses and most visualization^36^. The count matrix was filtered to remove cells with >10% of mitochondrial genes or low gene counts (<600 for enriched B cells, <200 for sorted B cells). The normalized data were integrated into one Seurat data file using the IntegrateData function. Principal component analysis was performed using variable genes. We compared the ranking of principle components (PCs) with the percentage of variance and determined the number of first ranked PCs used to perform Uniform Manifold Approximation And Projection (UMAP)^10^ to reduce the integrated dataset into two dimensions. Afterwards, the same number of first ranked PCs were used to construct a shared nearest-neighbor graph (SNN), and this SNN used to cluster the cells. For sorted cells, all sorted cells were used in the UMAP projection. For enriched B cells, immune cell clusters were identified using canonical markers. Contaminant cells (non-B cells) were removed and the data was normalized, integrated, and clustered with only the B cells. Specific B cell clusters were identified using canonical B cell markers^11^(Extended Data Fig. 1).

### Identification of differentially expressed genes (DEG) and functional enrichment

We performed differential gene expression testing using the FindMarkers function in Seurat with Wilcoxon rank sum test and the Benjamini–Hochberg method was used to adjust the p-values for multiple hypothesis testing. DEGs were filtered using a minimum log2(fold change) of 0.25 and a maximum FDR value of 0.05. Pathway analysis for the DEGs was conducted using the Metascape^35^.

### VDJ Sequence analysis

BCR VDJ regions were generated for each sample using the Cell Ranger Pipeline (v.6.0.1). BCR sequences were then filtered to include cells that have one light and one heavy chain per cell. Consensus sequences were aligned to germline variable-chain immunoglobulin sequences with IMGT HighV-QUEST v1.8.3^37^. Clonal expansions were defined based on sharing the same heavy and light chain V and J genes with >70% amino acid identity in heavy and light chain CDR3s.

### Recombinant monoclonal antibody (rmAb) production

Heavy chain and light chain variable sequences were codon optimized and cloned into in-house vectors, containing human IgG1 constant region or kappa or lambda constant regions, respectively. Expi293F cells were transfected with heavy chain and light chain plasmids using FectoPro (Polyplus transfection). Media was harvested after seven days, and rmAbs were purified with AmMag Protein A magnetic beads (Genscript,). Antibody concentrations were measured with a nanodrop spectrophotometer (Thermo Fisher Scientific) and human IgG quantitation ELISAs (Bethyl Laboratories).

### ELISA

For protein ELISAs, MaxiSorp 384-well plates (Thermo Fisher Scientific) were coated with 1 µg/ml recombinant SARS-CoV-2 S2 protein (Acro, S2N-C52H5), SARS-CoV-2 RBD (Acro, SPD-C52H3), or SARS-CoV-2 S1 (Acro, S1N-C52H3), HCoV-OC43 Spike protein (Sino Biological, 40607-V08B), HCoV-HKU1 Spike protein (Sino Biological, 40606-V08B), HCoV-229E Spike protein (Sino Biological, 40605-V08B), HCoV-NL63 Spike protein (Sino Biological, 40604-V08B) in carbonate-bicarbonate buffer at 4°C overnight. Plates were washed 6 times with PBST (PBS + 0.1% Tween20) after each step. The plates were blocked with blocking buffer (PBS + 1% BSA) for 1 hour at room temperature. Human plasma was serially diluted, and added for 1 hour at room temperature. Human rmAbs were added at concentrations of 10µg/ml and three 10-fold serial dilutions, and incubated overnight at 4°C. Secondary HRP-conjugated antibodies goat anti-human IgG (Bethyl Laboratories) or HRP-conjugated goat anti-human IgA (Bethyl Laboratories) were applied for 1h at RT, and plates were developed with TMB substrate (Thermo Fisher Scientific), and stopped with 2N sulfuric acid. Plates were read on a GloMax Explorer Microplate Reader (Promega).

### Bio-Layer Interferometry

rmAb interactions with S2, RBD, and S1 protein were measured on an Octet Red96e (Fortebio / Sartorius). Association and dissociation curves were measured with rmAbs bound to anti-human IgG Fc Capture (AHC) sensors at 20 nM and antigens in solution at 0, 16.7, 50, 150, and 450 nM in 1x kinetic buffer (Fortebio / Sartorius). BLI analysis software (Fortebio / Sartorius) was used for data processing and analysis. Buffer controls were subtracted from antigen values and curves were fitted globally for each group, consisting of all concentrations of the same ligand. Association and dissociation curves and constants as well as KD values for each antibody were reported and graphed with GraphPad Prism.

### Cell culture

Expi293F cells were cultured in 33% Expi293 Expression Medium (Gibco) and 67% Freestyle293 Expression Medium (Gibco). HeLa-ACE2 were kindly provided by of Dennis Burton^38^ and were cultured in Eagle’s Minimum Essential Medium (ATCC, 30-2003) with 10% heat-inactivated FBS (Corning) and 100 U/ml of penicillin–streptomycin (Gibco). Lenti X 293T cells (Takara Bio) were cultured in DMEM (ATCC, 30-2002) with 10% heat-inactivated FBS (Corning) and 100 U/ml of penicillin–streptomycin (Gibco).

### Generation SARS-CoV-2 spike pseudotyped lentiviral particles

Pseudotyped lentiviral particles were generated as previously described^39^. Briefly, LentiX 293T cells were seeded in 10cm plates. After 24 hours, cells were transfected using Fugene transfection reagent (Promega) with pHAGE-CMV-Luc2-IRES-ZsGreen-W, lentiviral helper plasmids (HDM-Hgpm2, HDM-tat1b, pRC-CMV-Rev1b), and wildtype or variant SARS-CoV-2 spike plasmids (parent plasmids publicly available from Jesse Bloom lab). After 48 to 60 hours, viral supernatants were collected and spun at 1000xg for 10m to remove cell debris. The lentiviral supernatants were concentrated using LentiX concentrator (Takara) according to the manufacturer’s instructions. The lentiviral pellets were resuspended at 20-fold viral increase in EMEM media and stored at -80°C. Virus was titrated on HeLa-ACE2 cells.

### Viral inhibition assays

Neutralization assays were performed as previously described^39^. Briefly, eight-fold serially diluted plasma starting at 1:80 from vaccinated individuals or five-fold serially diluted monoclonal antibodies starting at a concentration of 10µg/ml were incubated with SARS-CoV-2 pseudotyped virus for 1 hour at 37°C. The mixture was added to HeLa-ACE2 cells plated the prior day. After ∼50 hours post-infection, luciferase activity was measured on a GloMax Explorer Microplate Reader (Promega).

### Statistics and software

GraphPad Prism version 8.4.1 and R version 3.6.1 were used for statistical analyses. Statistical tests used and significance levels are indicated in the respective methods section or in the figure legends. Graphical illustrations were created with BioRender.

## Data Availability

The raw sequencing data will be uploaded in the GEO database before final publication.

## Acknowledgements

We thank all study participants who devoted time to our research. We thank Dennis Burton for providing the ACE2-HeLa cells. We thank Matthew Baker for key discussions. We thank Shaghayegh Jahanbani for antibody production. This work was supported by T32 AI007290-35 to R.C.B.; National Science Foundation Graduate Research Fellowship to R.C.B; the German Research Foundation (DFG, LA3657/1) to T.V.L.; NIH R01 AR063676 and U19 AI110491 to W.H.R.

## Author contributions

Author contributions: Conceptualization, R.C.B., T.V.L., W.H.R.; Methodology, R.C.B., T.V.L., N.S.R., L.J.L.; Software, R.C.B., T.V.L.; Validation, R.C.B., T.V.L.; Formal Analysis, R.C.B., T.V.L.; Investigation, R.C.B., T.V.L.; Resources, R.C.B., T.V.L., W.H.R.; Data Curation, R.C.B., T.V.L.; Writing – Original Draft, R.C.B., T.V.L.; Writing – Review & Editing, R.C.B., T.V.L., N.S.R., L.J.L., W.H.R.; Visualization, R.C.B., T.V.L.; Supervision, T.V.L., W.H.R.; Project Administration, R.C.B., T.V.L., W.H.R.; Funding Acquisition, R.C.B., T.V.L., W.H.R..

## Materials Availability

Materials generated in this study will be made available on request and may require a material transfer agreement.

## Data Availability Statements

The raw sequencing data will be uploaded in the GEO database before final publication.

---

**Extended Data Figure 1:**
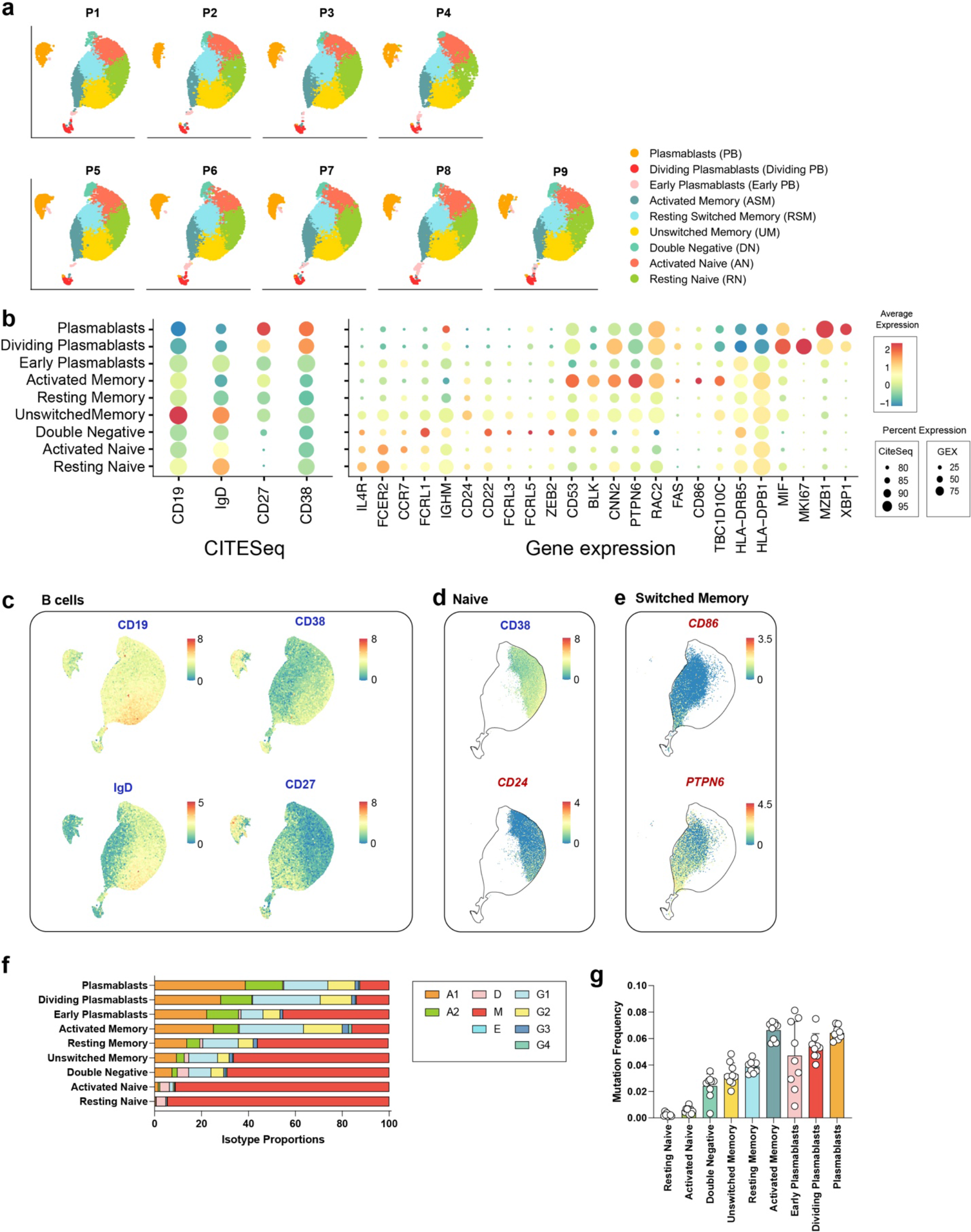
Classification of UMAP B cell clusters. **a**, Distribution of B cell clusters for each individual (n=9) included in the study. **b**, Gene and cell surface expression of B cell clusters, represented as heatmap / dot plots. **c**, UMAP embedding of B cells colored by CD19, CD38, IgD, and CD27 protein expression (blue). **d**, UMAP embedding of naïve B cells colored by CD38 protein expression (blue), and *CD24* gene expression (red). **e**, UMAP embedding of memory B cells colored by *CD86* and *PTPN6* gene expression (red). **f**, Average proportions of isotype subclasses in each B cell cluster in *n*=9 individuals vaccinated individuals across all timepoints. **g**, Average combined mutation frequency (number of mutations per length of V-gene) in light chain and heavy chain V-gene regions in *n*=9 individuals in the respective nine different B cell clusters. Individual values, means, and standard deviations are shown.

**Extended Data Figure 2:**
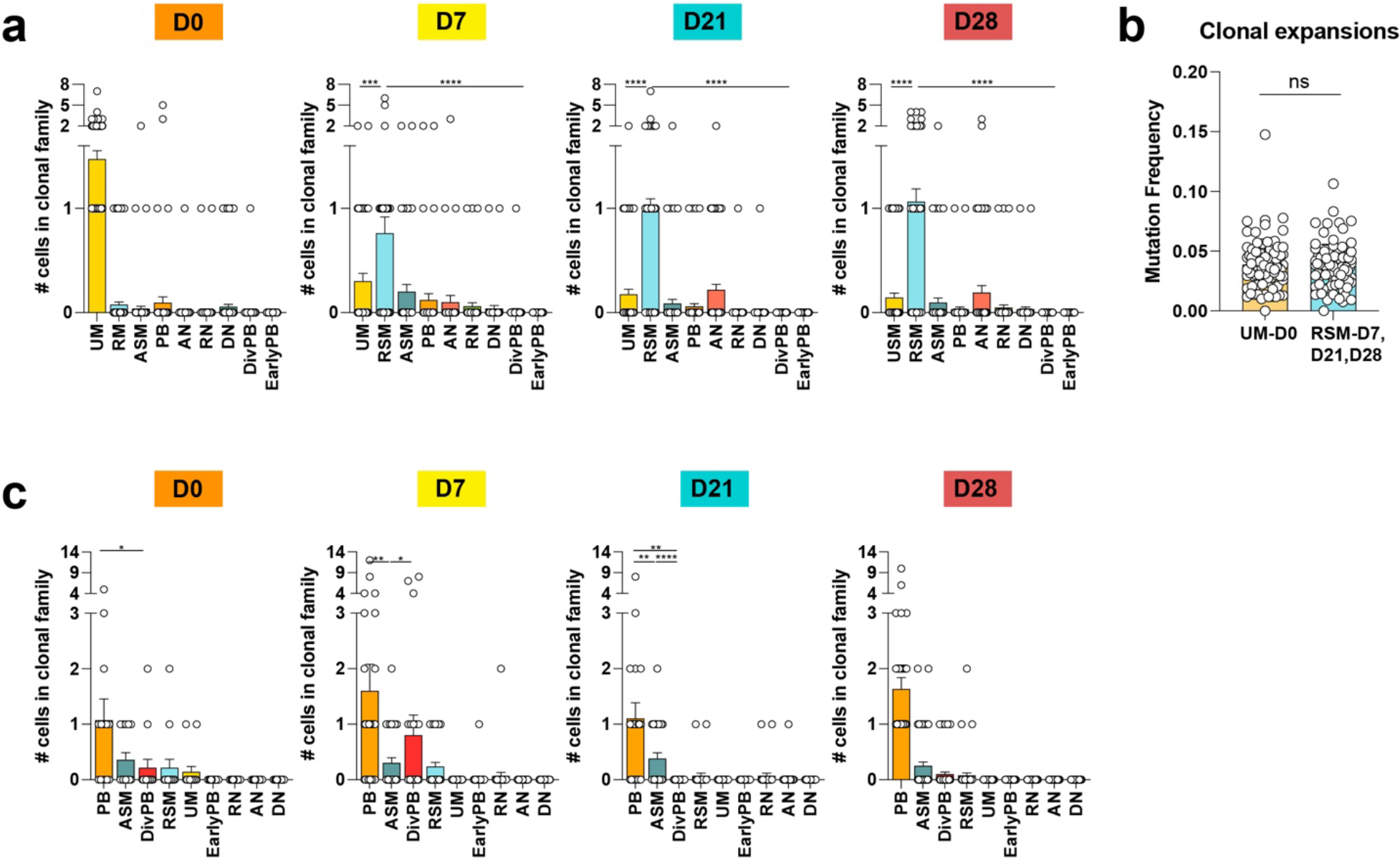
Characterization of clonal B cell expansions in response to mRNA vaccination. **a**, Cells corresponding to clonal expansions shown in (Fig 2a). Number of cells in each clonal expansion (*n*=106) in each B cell cluster at all four timepoints. Statistics are not calculated for D0 as each clonal family is set to contain at least one clone at D0 in the UM B cell cluster. **b**, Average mutation frequency (number of mutations per length of V-gene) of clonal UM expansions at D0 and clonal RSM expansions at D7, D21, and D28. Each dot represents the average of each mutation frequency for the clonal family in specified B cell clusters and time. **c**, Cells corresponding to the clonal expansions shown in (Fig. 2d). Number of cells in each clonal expansion (*n*=52) in each B cell cluster at all four timepoints. Statistics are not calculated for D28 as each clonal family is set to contain at least one clone at D28 in the PB cluster. Individual values, and **a,c**, standard error of the means, and **b**, standard deviations are shown. \*\**P* <0.01, \*\*\**P* <0.001, \*\*\*\**P* <0.0001 according to **a,c**, two-tailed Kruskal-Wallis test and **b**, paired two-tailed t test.

**Extended Data Figure 3:**
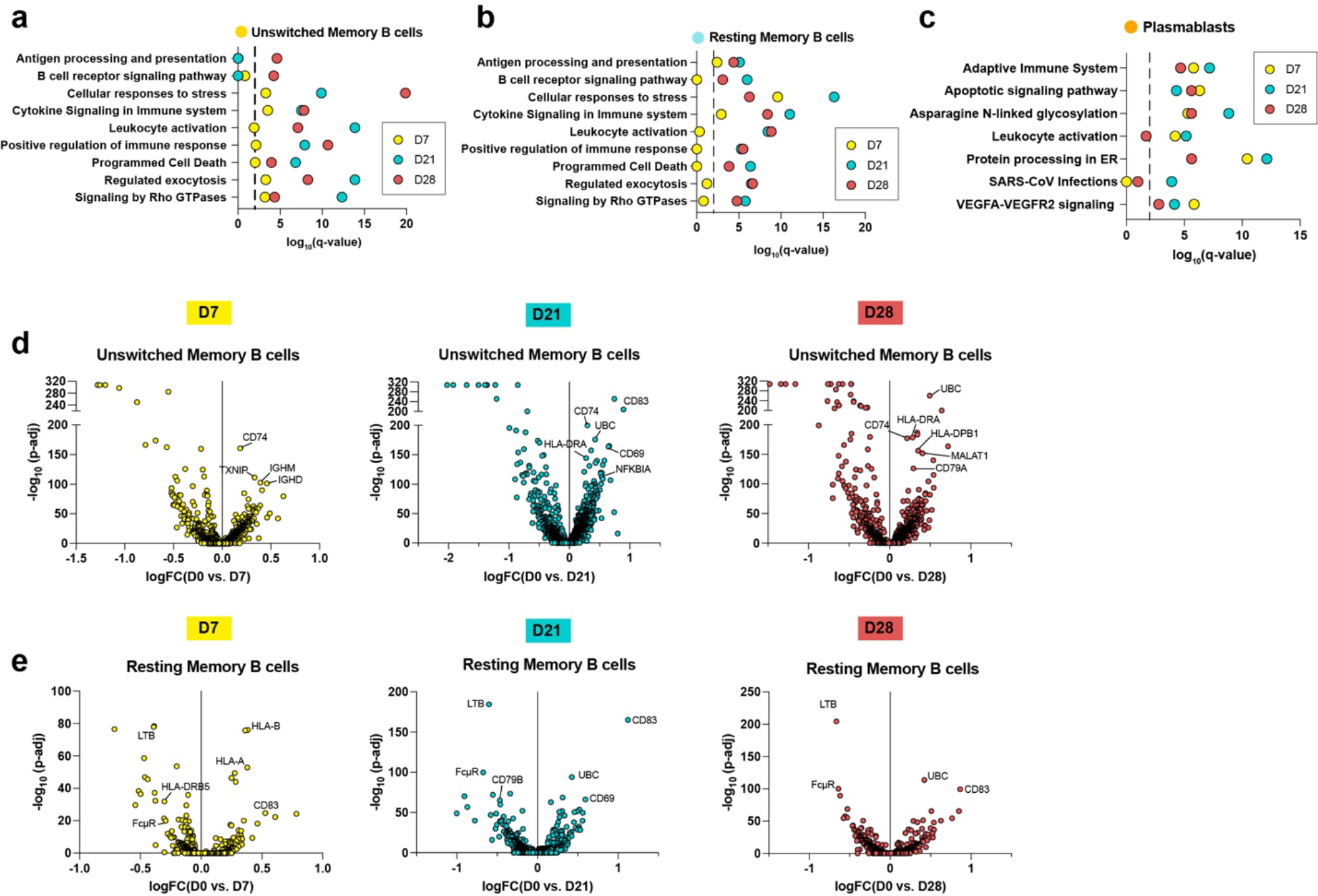
Pathway enrichments and differentially expressed genes in response to mRNA vaccination. **a-c**, Gene enrichment signatures of DEGs via Metascape^35^ in **a**, UM, **b**, RSM, and **c**, PB for D7 (yellow dots), D21 (turquoise dots), D28 (red dots) versus pre- vaccination. Dashed line represents q < 0.01. **d,e**, Volcano plot of differentially expressed genes in **d**, USM and **e**, RSM at timepoints D7 (left), D21 (center), and D28 (right) relative to D0.

**Extended Data Figure 4:**
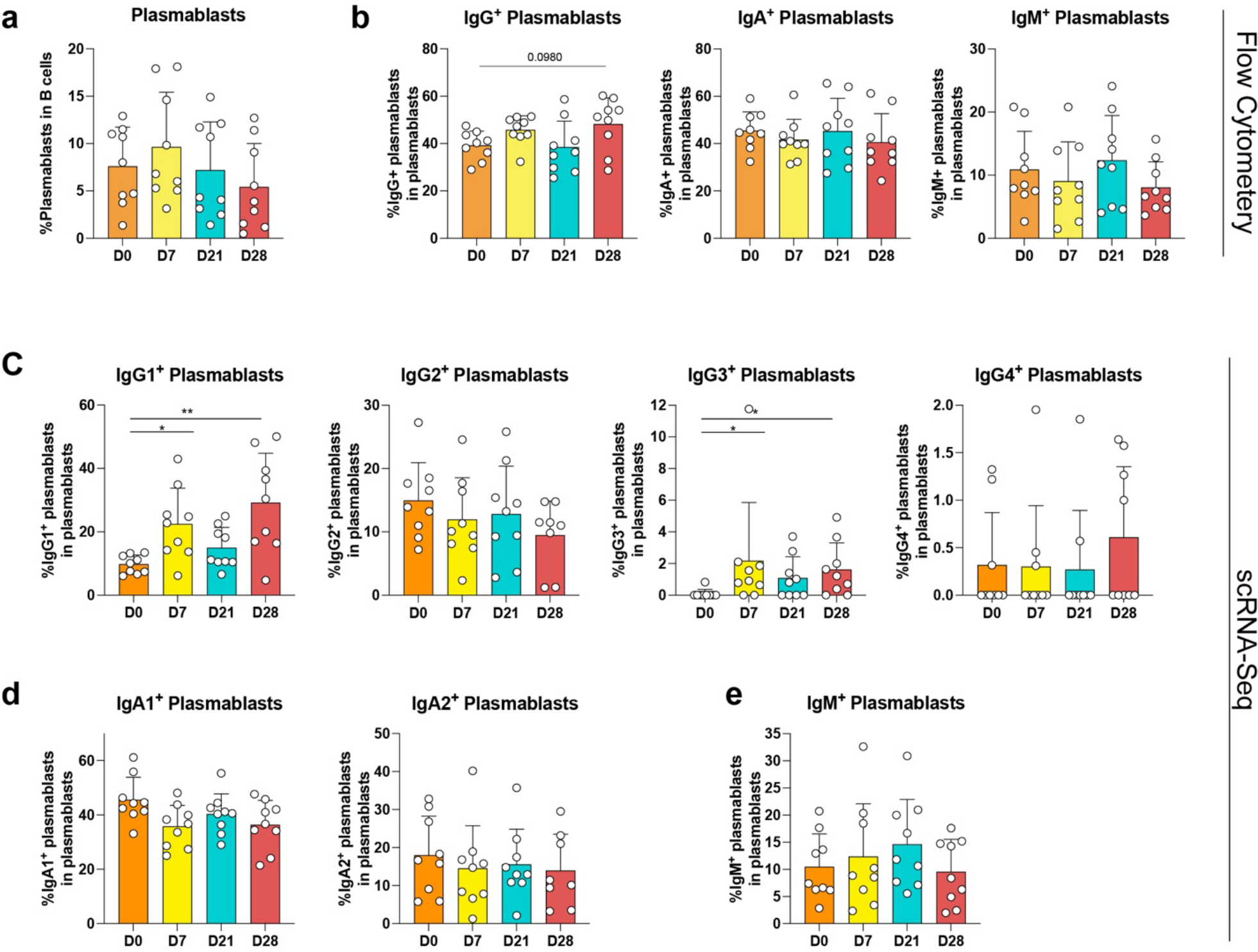
Global PB proportions measured by flow cytometry and scRNA-Seq. **a,b**, Flow cytometry data, **a**, mean percentages of PB in all B cells at the four indicated timepoints. **b**, Proportions of IgG^+^ (left), IgA^+^ (center), and IgM^+^ PB (right) of all PB. **c-e**, scRNA-Seq data, proportions of **c**, IgG subclasses, **d**, IgA subclasses, and **e**, IgM in total PB. n=9 individual values, means, standard deviations are shown. \**P* <0.05, \*\**P* < 0.01, \*\*\**P* < 0.001, \*\*\*\**P* < 0.0001, or exact value according to two-tailed one-way ANOVA test.

**Extended Data Figure 5:**
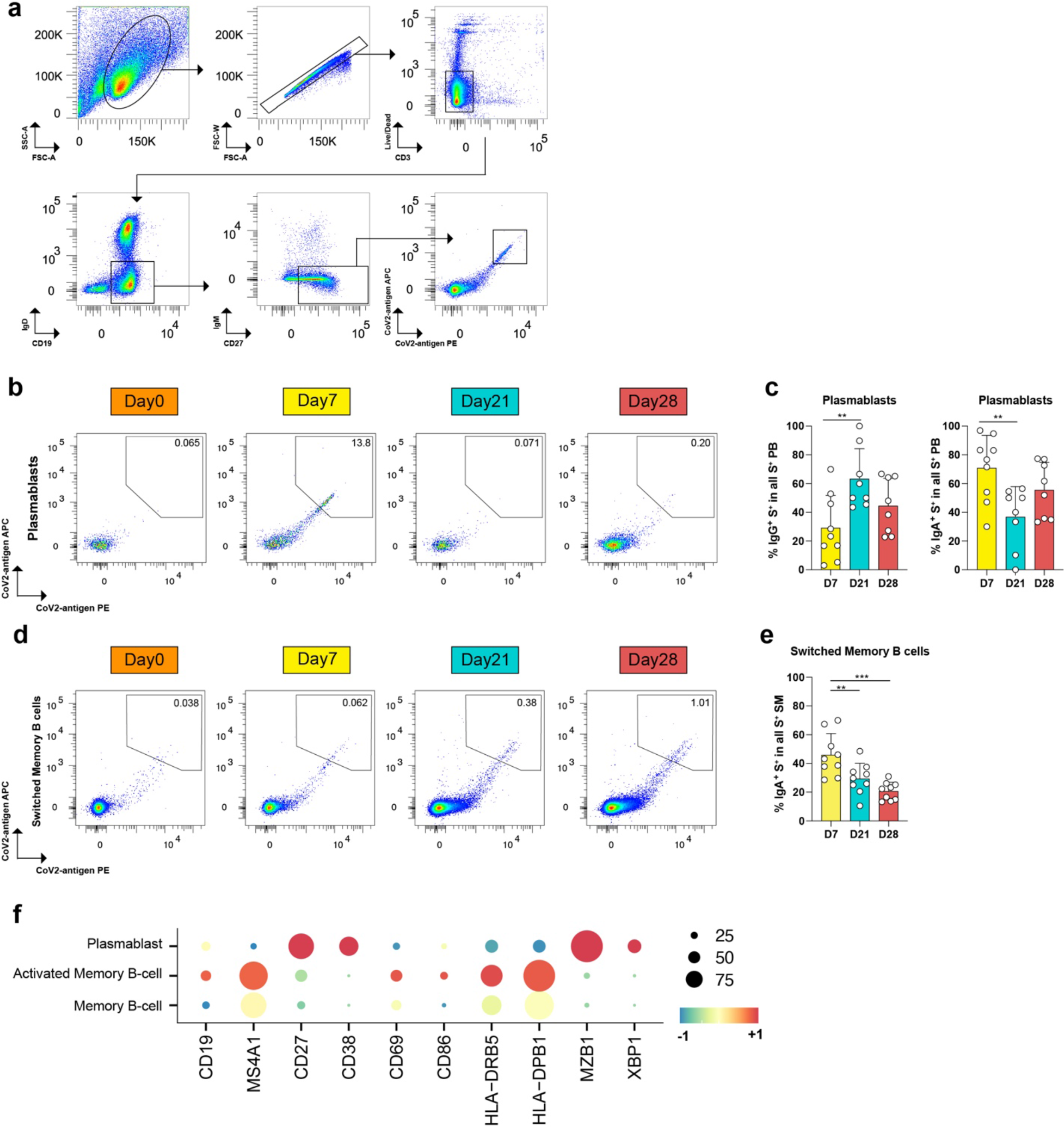
Characterization of sorted SARS-CoV-2^+^ SM B cells and PB. **a**, Gating strategy for sorting on live singlet SM and PB based on marker expression: CD3^-^, Cd19^+^, IgD^-^, IgM^-^, CD27^+^. Cells were stained with two antigen-tetramers for each subunit of S (S1, S2, and RBD), respectively, labeled with PE and APC. Double-positive cells were sorted. **b**, Representative flow cytometry plots of S+ PB. **c**, Proportion of IgA^+^ (left) and IgG^+^ (right) S^+^ PB in all S^+^ PB at the indicated timepoints post vaccination. **d**, Representative flow cytometry plots of S^+^ SM. **e**, Proportion of IgA^+^ S^+^ SM in all S^+^ SM at the indicated timepoints post vaccination. **f**, Dot plots of gene expression of B cell clusters shown in (Fig. 3d). **c,e**, n=9 individual values, means, and standard deviations are shown. \**P* <0.05, \*\**P* < 0.01, \*\*\**P* < 0.001 according to two-tailed one-way ANOVA test.

**Extended Data Figure 6:**
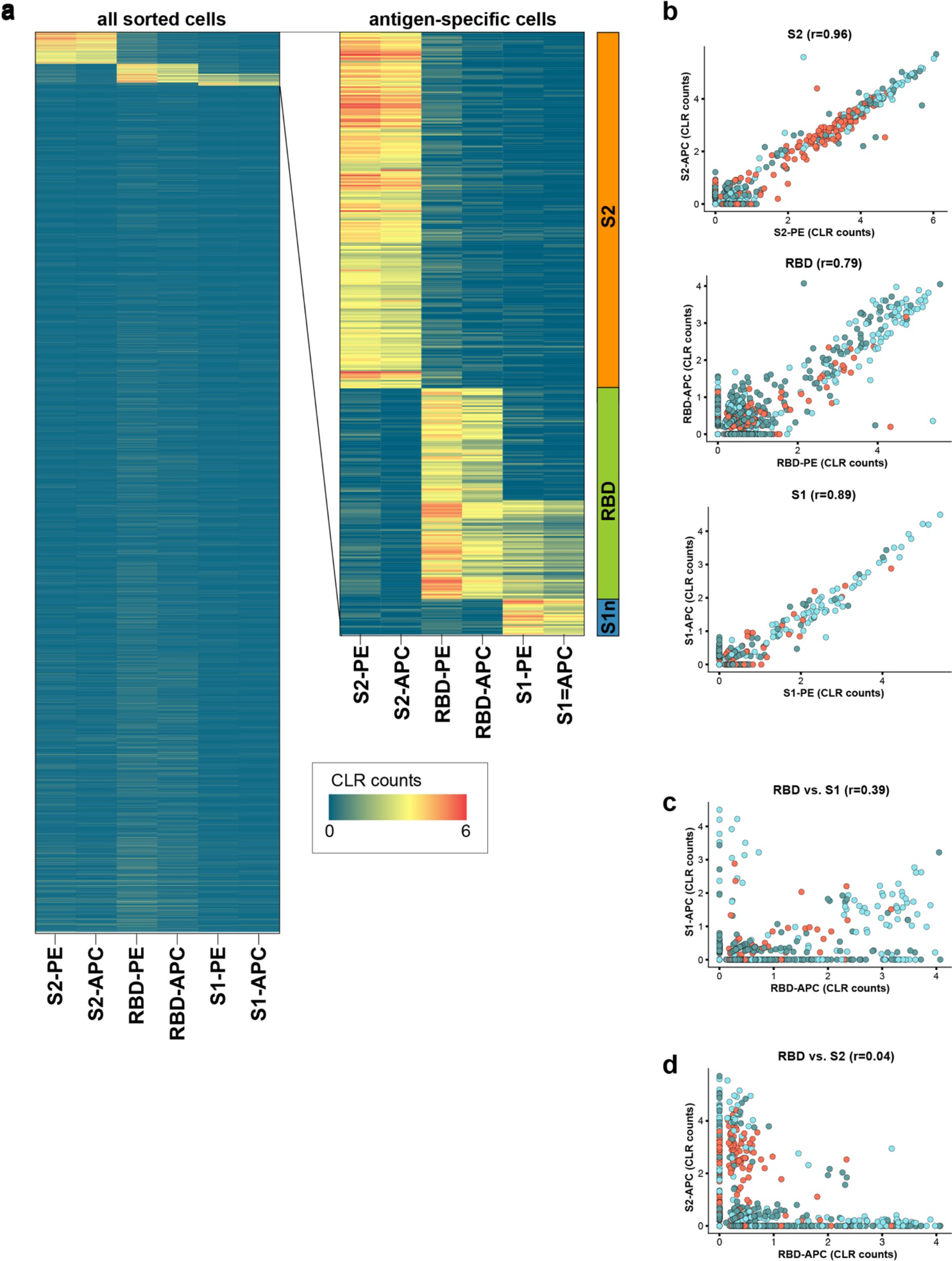
Specific binding of S antigen tetramers to sorted cells. **a-d**, Single-cell sequencing data analysis of barcodes in de-multiplexed data set. Antigen-positive and negative CD3^-^, Cd19^+^, IgD^-^, IgM^-^, CD27^+^ B cells were mixed during the sort. **a**, Heatmap of centered log ratio transformed (CLR) counts of barcoded PE and APC tetramers with S2, RBD, and S1 for all sorted cells. Inset magnifies the antigen-specific cells used in the analysis. **b**, Correlation of barcoded PE and APC tetramers S2 (top), RBD (middle), and S1 (bottom). **c**, Correlation of barcoded APC tetramers of S1 and RBD. **d**, Correlation of barcoded APC tetramers of S2 and RBD.

**Extended Data Figure 7:**
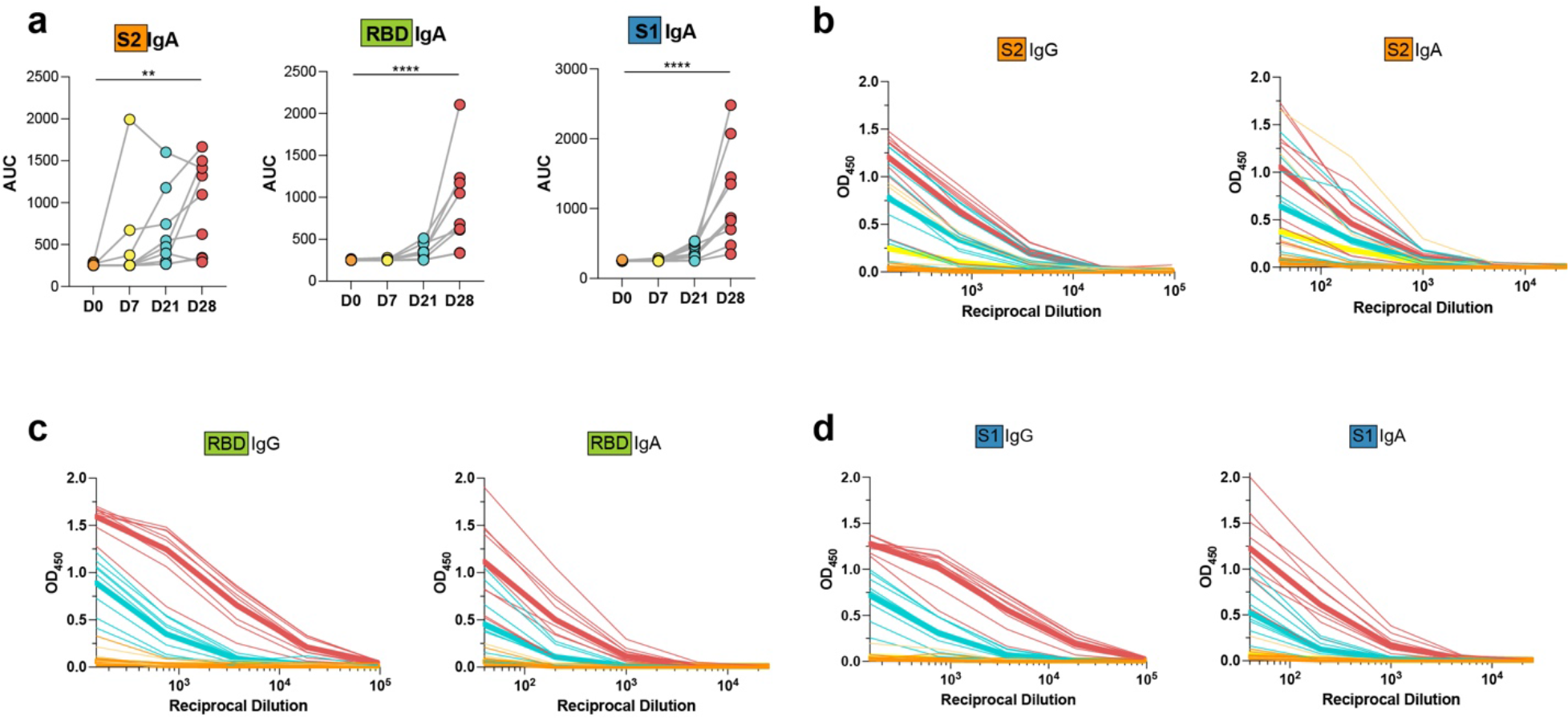
Plasma reactivity to S protein subunits. **a**, Area under the curve (AUC) for anti-S2 IgA (left), anti-RBD IgA (center), and anti-S1 IgA (right) from n=9 vaccinated individual plasma samples. **b-d,** Plasma IgG (left) and IgA (right) reactivity to **a**, S2, **b**, RBD, **c**, S1 as determined by ELISA. Graph shows optical density units at 450nm of serial plasma dilutions. \**P* <0.05, \*\**P* < 0.01, \*\*\**P* < 0.001, according to two-tailed one-way ANOVA test.

**Extended Data Figure 8:**
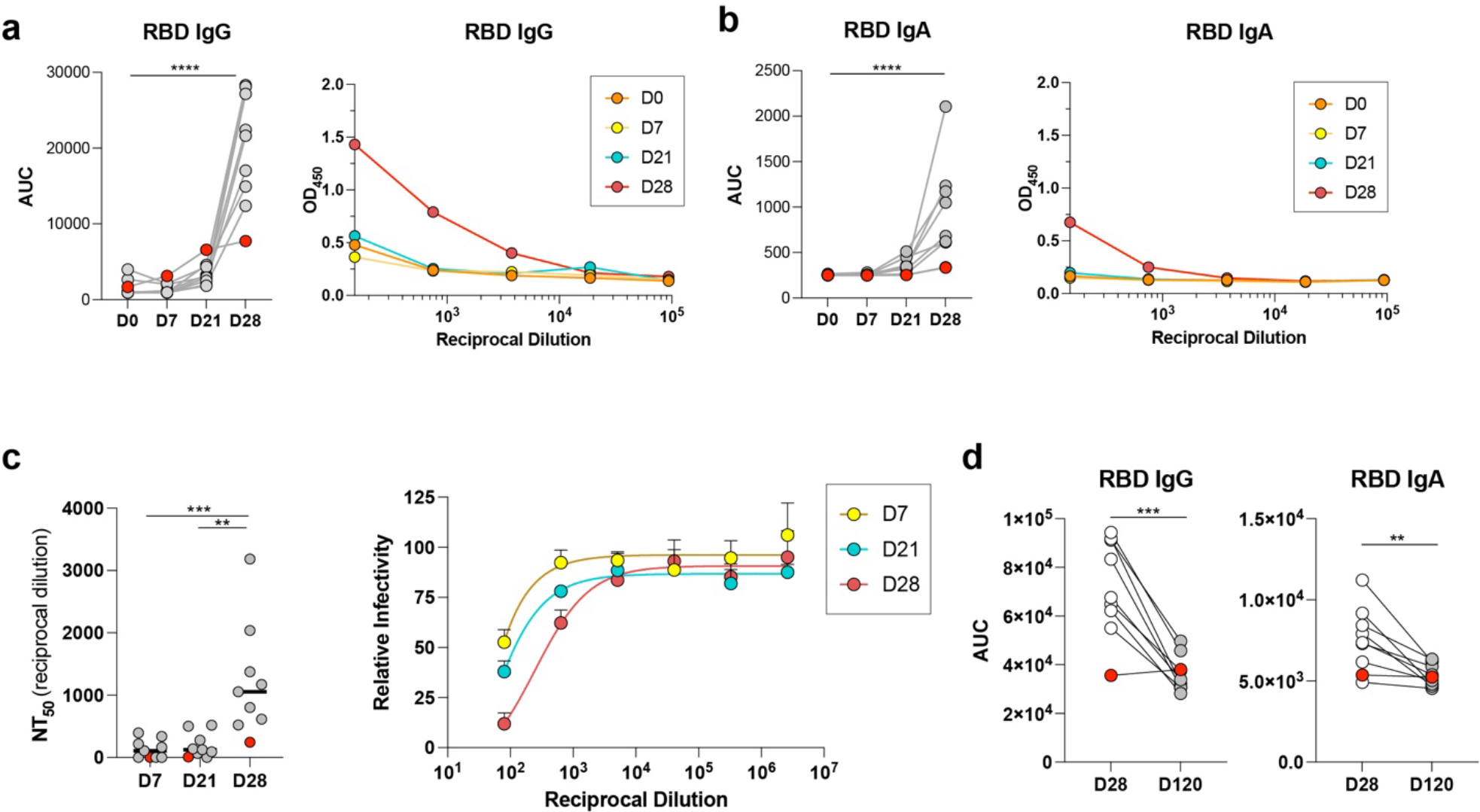
Analysis of individual P1 who contracted COVID-19 eight weeks post second vaccination. **a-b**, Plasma reactivities of **a**, IgG and **b**, IgA to RBD as measured by ELISA in serial plasma dilutions (right) and depicted as AUC (left). **c**, Plasma neutralization titers (left) and neutralization curves for plasma of individual P1 at D7, D21, D28 (right). **d**, Plasma reactivities to RBD for IgG (left) and IgA (right), on D28 compared to day 120 (D120), two weeks after COVID-19 diagnosis. P1 is highlighted in red, the other eight individuals are grey. \**P* <0.05, \*\**P* < 0.01, \*\*\**P* < 0.001 according to **a-c**, two-tailed one-way ANOVA test and **d**, paired t-test.

**Extended Data Figure 9:**
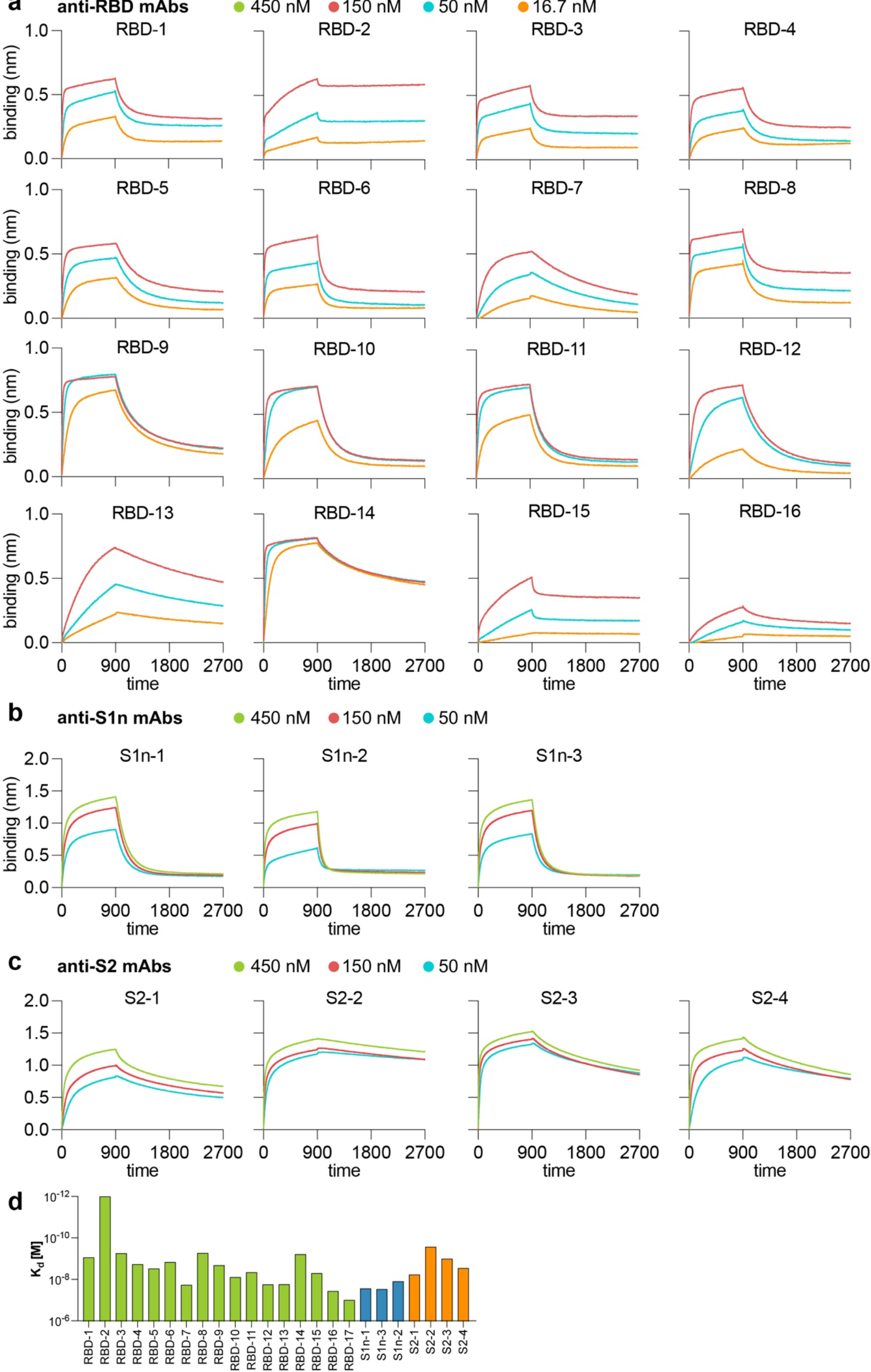
Binding affinities of anti-RBD, S1n, and S2 rmAbs. **a-c**, rmAb-antigen interaction intensities were measured by bio-layer interferometry with immobilized onto Octet sensors, and S protein subunits. **a,** RBD, **b**, S1n, and **c**, S2 in solution. Binding curves are shown for the indicated concentrations of antigen. **d**, Kd values of antibodies shown in a-c.

**Extended Data Figure 10:**
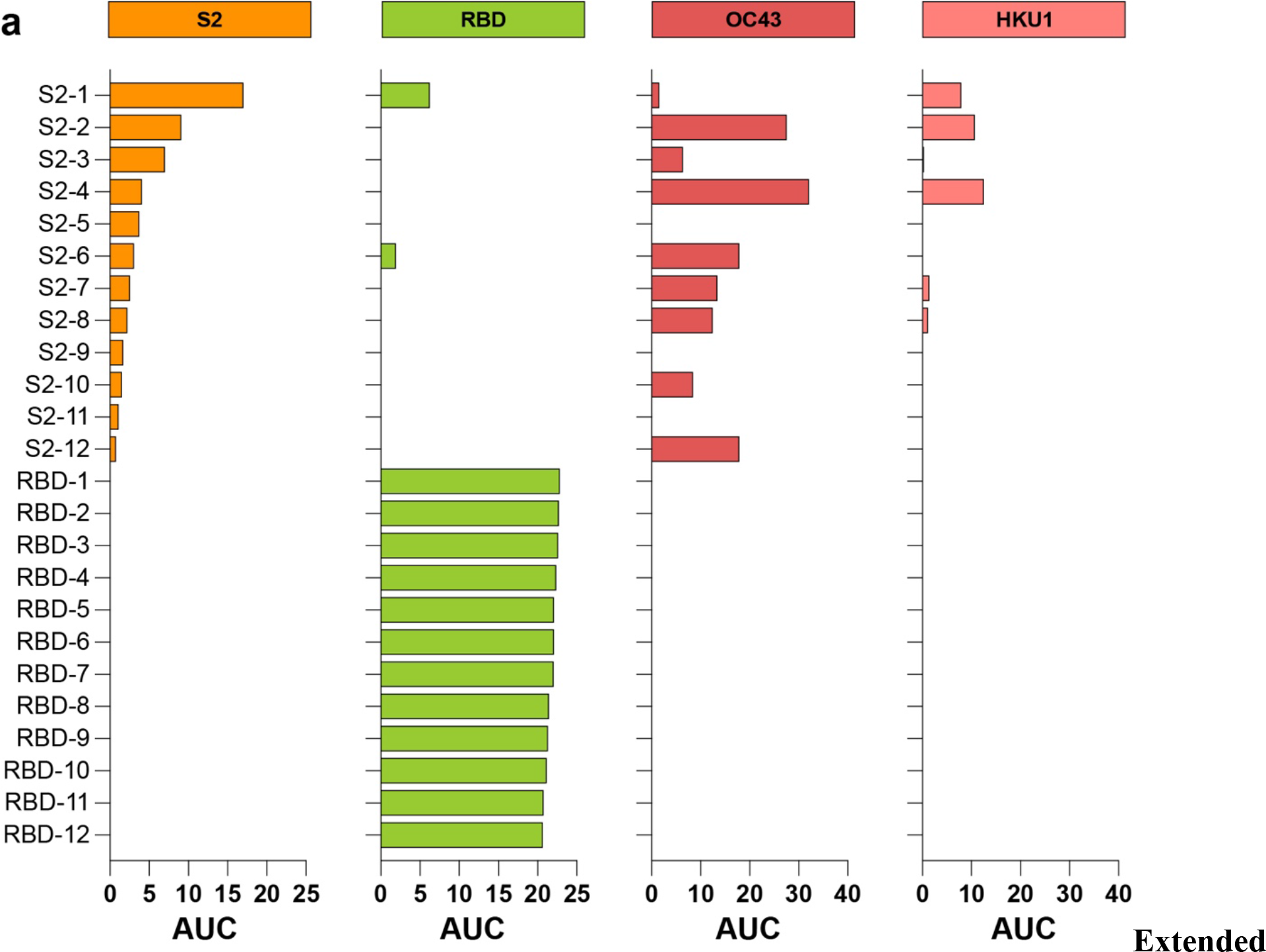
Anti-S2 rmAbs are cross-reactive to other human-pathogenic beta-coronavirus S proteins. **a**, Binding of anti-S2 and anti-RBD rmAbs to SARS-CoV-2 S2, SARS-CoV-2 RBD, HCoV-OC43 spike, and HCoV-HKU1 spike, assessed by ELISA and represented as AUC of optical density measurements of serial dilutions. No binding was observed for HCoV-229E and HCoV-NL63 spike proteins. Threshold, represented as 0, was set to the average binding to BSA plus three times the standard deviation of background binding to BSA.

**Extended Data Figure 11:**
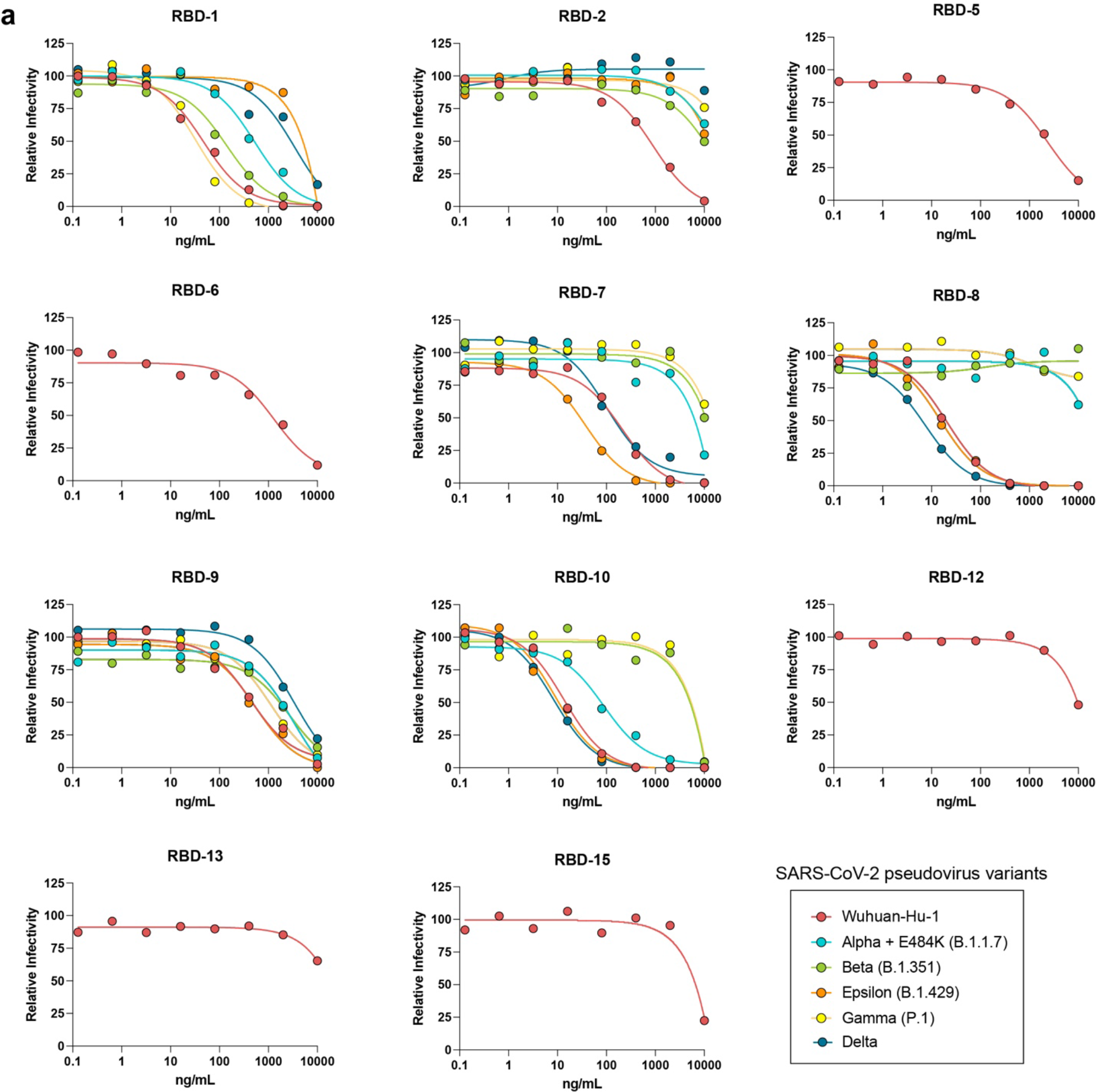
Neutralization of SARS-CoV-2 Wuhan-Hu-1 pseudovirus and variants by rmAbs from vaccinated individuals. **a**, Neutralization curves for each selected rmAb against the five strains, measured by luminescence measurement upon SARS-CoV-2 pseudoviruses infection of transgenic HeLa cells expressing human ACE2. Pseudovirus strains: Wuhan-Hu-1, Alpha+E484K, Beta, Gamma, Epsilon, and Delta.

**Extended Data Table 1.** Characteristics of participants in BNT162b2 vaccination study.

| Participant | Age | Sex | Race | Ethnicity | Symptoms of BNT162b2 |  | Site of vaccination |  | Confirmed SARS-CoV-2 infection |  | Symptoms to SARS-CoV-2 infection |
| --- | --- | --- | --- | --- | --- | --- | --- | --- | --- | --- | --- |
|  |  |  |  |  | Dose 1 | Dose 2 | Dose 1 (site) | Dose 2 (site) | Prior | Post |  |
| P1 | 36-40 | M | Caucasian | Hispanic | Site tenderness | Site tenderness | Left | Left | No | Yes | Loss of smell, muscle aches, and fatigue |
| P2 | 21-25 | M | Asian | Not Hispanic | Site tenderness | Fever, muscle aches, fatigue, chills | Left | Left | No | No | N/A |
| P3 | 31-35 | M | Asian | Not Hispanic | Site tenderness, muscle aches | Site tenderness, muscle aches, headache, chills | Left | Left | No | No | N/A |
| P4 | 26-30 | M | Asian | Not Hispanic | None | Site tenderness, fever, muscle aches, fatigue, chills, loss of appetite | Left | Left | No | No | N/A |
| P5 | 31-35 | F | Asian | Not Hispanic | Site tenderness | Site tenderness, muscle aches, fatigue, headache | Right | Right | No | No | N/A |
| P6 | 46-50 | F | Caucasian | Not Hispanic | Site tenderness, site swelling | Site tenderness | Left | Left | No | No | N/A |
| P7 | 51-55 | F | Asian | Not Hispanic | Site tenderness, muscle aches, fatigue, headache, joint pain | Site tenderness, muscle aches, fatigue, headache, joint pain, fever, chills | Right | Right | No | No | N/A |
| P8 | 26-30 | F | Caucasian | Not Hispanic | Site tenderness | Site tenderness, site swelling, muscle aches, fatigue, headache | Left | Left | No | No | N/A |
| P9 | 36-40 | M | Caucasian | Not Hispanic | None | None | Right | Right | No | No | N/A |

**Extended Data Table 2.** SARS-CoV-2 pseudovirus variants used in neutralization assays.

| variant name | greek alphabet name | Classification | origin | mutations | RBD mutations (319-541) |
| --- | --- | --- | --- | --- | --- |
| **B.1.1.7 + 484K** | **Alpha + 484K** | **Variant of Concern** | UK | Δ69/70, Δ144, E484K, N501Y, A570D, D614G, P681H, T716I, S982A, D1118H | E484K, N501Y |
| **B.1.351** | **Beta** | **Variant of Concern** | South Africa | D80A, D215G, Δ241/242/243, K417N, E484K, N501Y, D614G, A701V | K417N, E484K, N501Y, |
| **P1** | **Gamma** | **Variant of Concern** | Japan / Brazil | L18F, T20N, P26S, D138Y, R190S, K417T, E484K, N501Y, D614G, H655Y, T1027I | K417T, E484K, N501Y, |
| **B.1.617.2** | **Delta** | **Variant of Concern** | India | T19R, T95I, G142D, Δ156/157, R158G, L452R, T478K, D614G, P681R, D950N | L452R, T478K |
| **B.1.429** | **Epsilon** | **Variant of Interest** | US / California | S13I, W152C, L452R, D614G | L452R |

**Extended Data Table 3.**
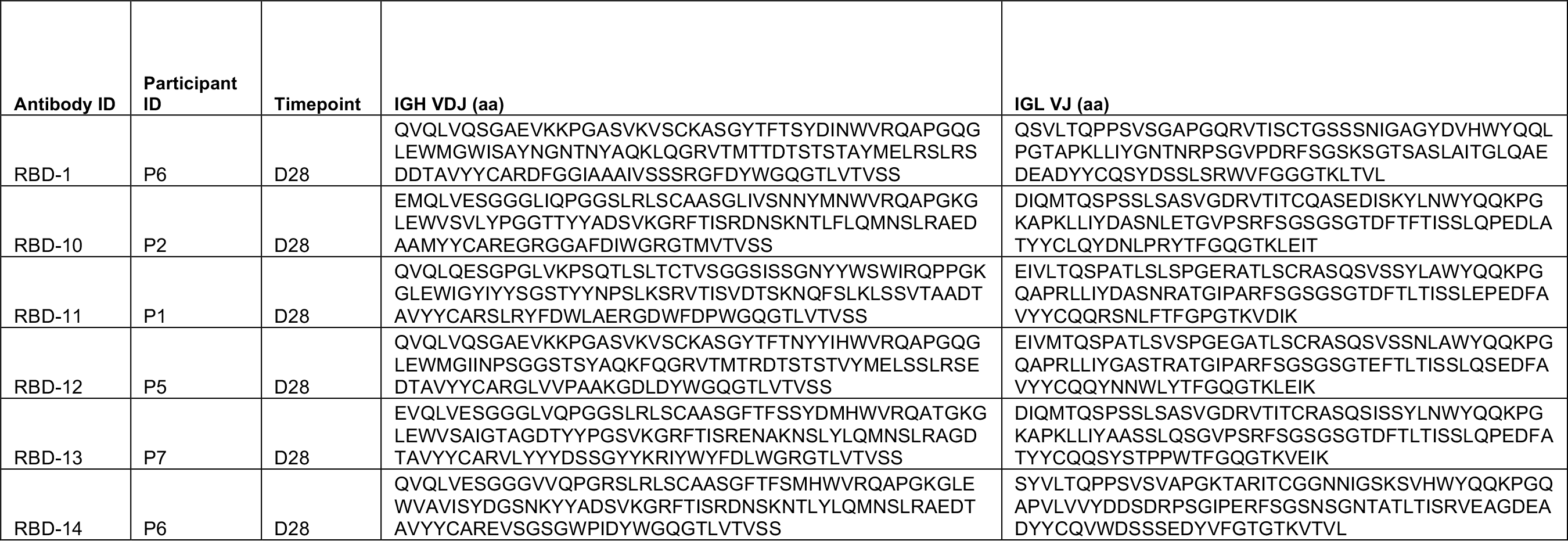
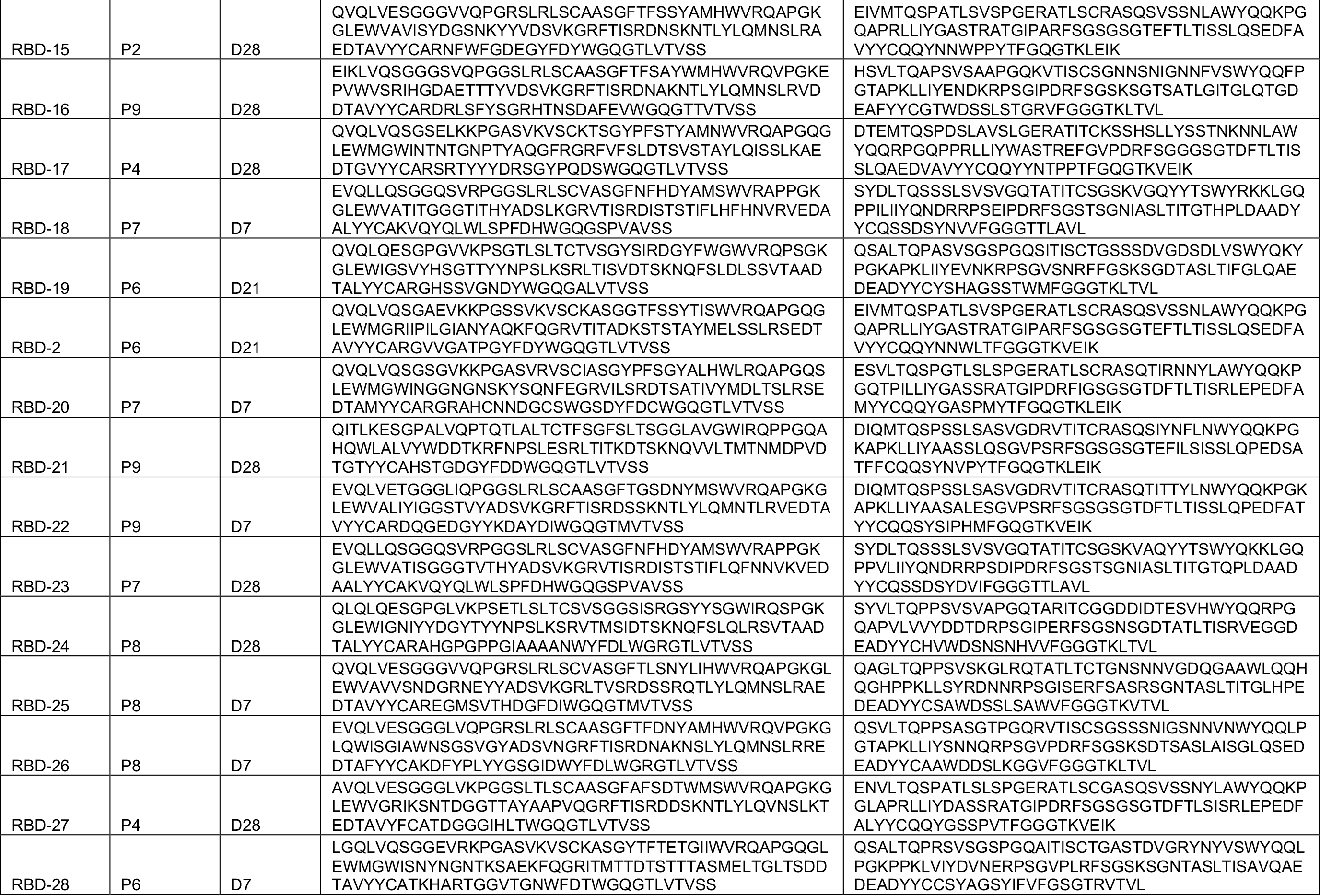
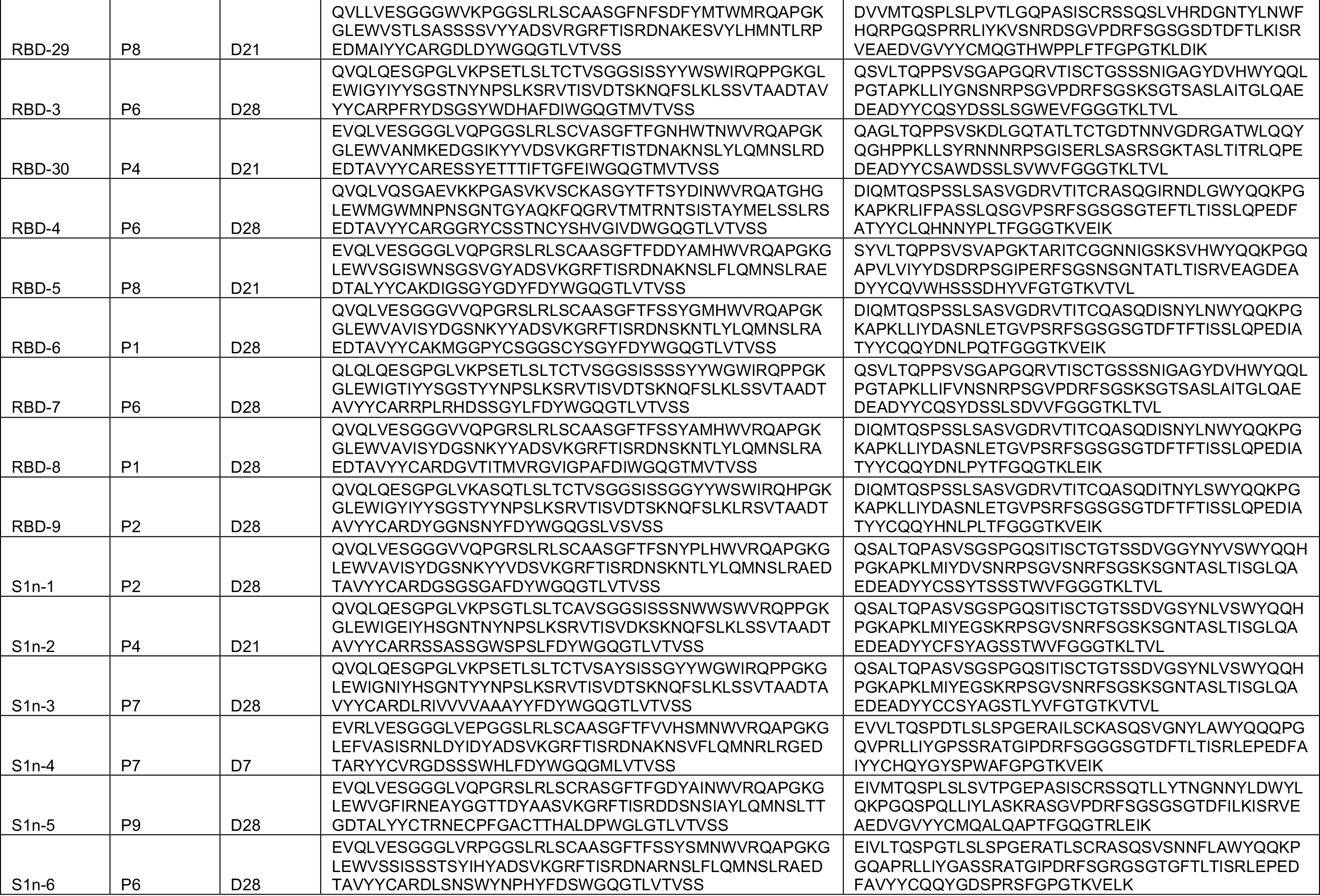
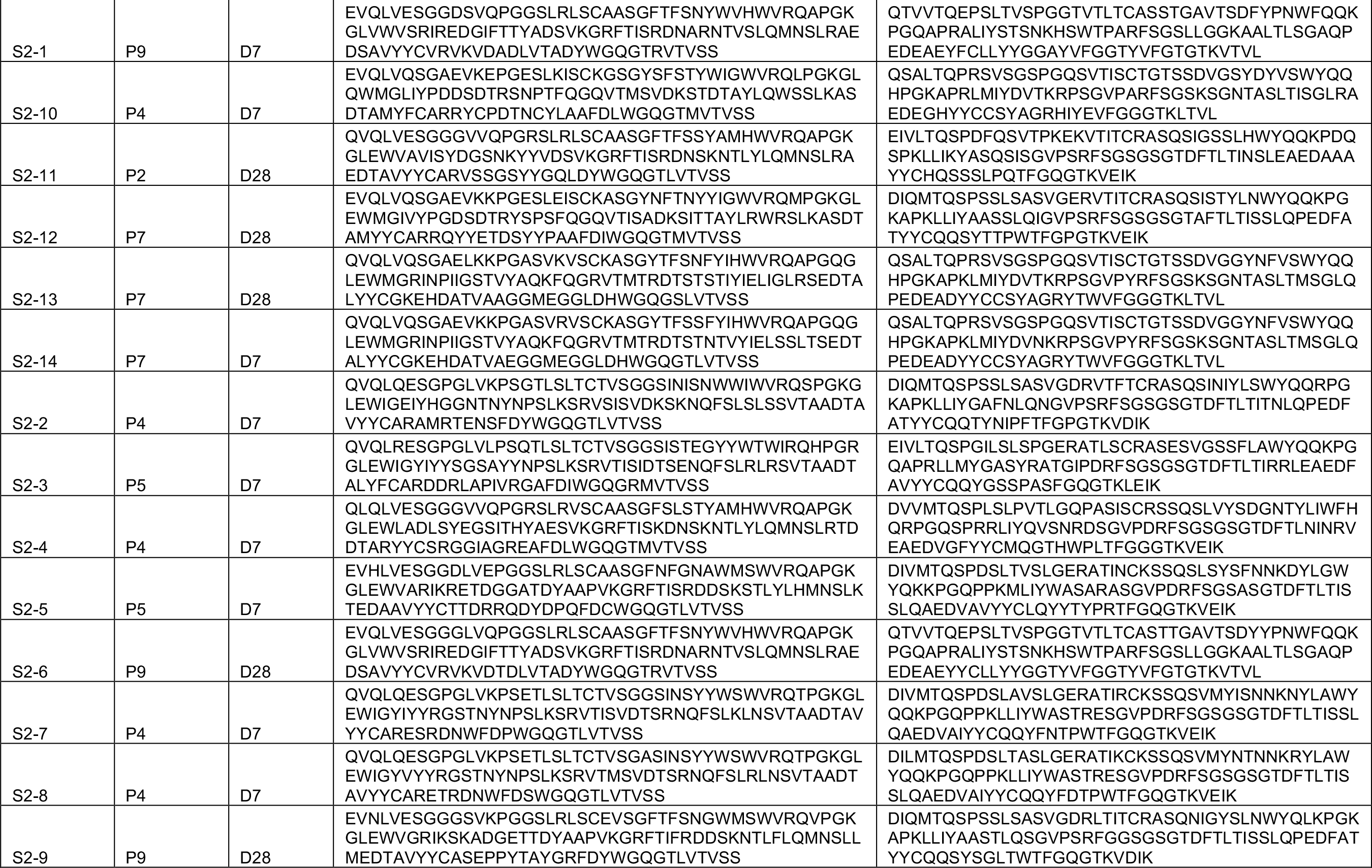
Expressed rmAbs derived from S^+^ sorted B cells

